# INCIDENCE OF VENOUS THROMBOTIC EVENTS AND EVENTS OF SPECIAL INTEREST IN A RETROSPECTIVE COHORT OF COMMERCIALLY-INSURED US PATIENTS

**DOI:** 10.1101/2021.06.13.21258854

**Authors:** Susan C. Weller, Laura Porterfield, John W. Davis, Gregg S. Wilkinson, Lu Chen, Jacques Baillargeon

## Abstract

**Objective:** To estimate the US incidence of thrombotic events and related rare diagnoses.

**Design:** Claims-based retrospective cohort study of incidence.

**Setting:** US commercial health insurance administrative claims database.

**Participants:** Adults 25-65 years of age between 2015 and 2019 with a minimum of 12 consecutive thrombosis-free months of continuous enrollment beginning 2014 were selected.

**Main Outcomes:** Age (10-year intervals) and sex stratum specific incidence rates per 100,000 person-years were determined for: venous thromboembolism (VTE), cerebral venous thrombosis (CVT), and other major venous thrombotic events, and events of special interest, including immune thrombocytopenic purpura (ITP), hemolytic-uremic syndrome (HUS), and heparin-induced thrombocytopenia (HIT).

**Results:** Of 13,249,229 enrollees (half female/male), incidence of venous thromboembolic events (DVT, PE, CVT, or other major venous thrombotic conditions) was 247.89 per 100,000 person-years (95% CI: 245.96, 249.84). Incidence of VTE was 213.79 with ICD codes alone (95% CI: 211.99, 215.59) and 127.18 (95% CI: 125.80, 128.58) when also requiring a filled anticoagulation prescription. Incidence was 6.37 for CVT (95% CI: 6.07, 6.69), 26.06 for ITP (95% CI: 25.44, 26.78), 0.94 for HUS (95% CI: 0.82, 1.06), and 4.82 for HIT (95% CI: 4.56, 5.10). The co-occurrence of CVT with either ITP or HIT (diagnoses within 14 days of one another) was 0.090 (95% CI: 0.06, 0.13). Incidence tended to increase with age and was higher for women under 55. Incidence for CVT, HUS, and CVT with ITP or HIT was higher for women in all age groups. Incidence of PE and CVT increased significantly over the five-year period, while DVT rates decreased.

**Conclusions:** These results are the first US estimates for incidence of thrombotic and rare events of interest in a large, commercially-insured US population. Findings provide a critically important reference for determining excess morbidity associated with COVID-19 and more generally for vaccine pharmacovigilance.

**BOX POINTS:** *What is already known on this topic?:*

- Incidence of venous thromboembolic diagnoses vary by country and date.
- There have been improvements in the past decade in thromboprophylaxis (e.g., for deep vein thrombosis) and detection (e.g., for cerebral venous thrombosis), but there are no recent comprehensive estimates for the United States.

*What this study adds:*

- Our results document the US incidence of thrombotic and related rare diagnoses for the most recent five-year pre-pandemic period (2015-2019) in an insured population.

---

Although interest in incidence of rare thrombotic events (e.g., cerebral venous thrombosis, CVT) has increased with recent SARS-CoV-2 virus (COVID-19) and vaccine adverse event surveillance,[1–3] few recent estimates exist for the United States. This study estimates the pre-pandemic incidence of thrombotic events and rare conditions associated with thrombocytopenia that are potentially associated with COVID-19 disease and/or vaccination. Using a large, US commercial healthcare administrative database, we provide the first estimates of incidence since improvements over the past decade in prevention and detection of thromboses. These data will be useful in evaluating morbidity associated with COVID-19 disease and more generally, for pharmacovigilance.

Incidence estimates for thrombotic events have been inconsistent and limited by population size. The focus has been on the more common diagnosis of venous thromboembolism (VTE), lower extremity deep vein thrombosis (DVT) and/or pulmonary embolism (PE), but estimates are highly variable across populations.[4–6] Limited data are available on the incidence of rarer thromboses because samples were too small to detect events. Recently, a report from Denmark[5] estimated combined incidence for major venous thromboses, as well as for VTE alone, and data from the European Union (EU) and the United Kingdom (UK)[6] estimated CVT incidence.

In this study, incidence of VTE, CVT, other thromboses, immune thrombocytopenic purpura (ITP), hemolytic-uremic syndrome (HUS), and heparin-induced thrombocytopenia (HIT) was estimated using administrative data from a large, US commercial health insurance program. Incidence was estimated stratified by sex and age, among adults aged 26-65 for the most recent five-year period prior to the pandemic (2015-2019).

## Methods

Diagnoses were identified using the Optum Clinformatics® Data Mart Database, one of the largest commercial insurance populations in the United States. The database contains de-identified inpatient and outpatient claims (medical, pharmaceutical, and laboratory) for approximately 62 million unique enrollees (2007-2018); 81% of whom have at least one medical claim per year and almost half of whom have continuous enrollment for two or more years. The distribution of age (25-64) and sex in the claims population is similar to that of the US population (within 1-3%), but more members reside in the southern US region (11% higher).[7]

Diagnoses were identified for: acute VTE (DVT: ICD-9: 451.1x, 451.2x, 451.81, 451.83; ICD-10: I80.1x, I80.2x, I80.3, I80.219; PE: ICD-9: 415.1x, ICD-10: I26.0x, I26.9x); CVT (ICD-9: 325.0, 437.6; ICD-10: G08, I67.6, I63.6) and other major venous thrombotic conditions, excluding pregnancy-related conditions (portal vein thrombosis, hepatic vein thrombosis/Budd-Chiari syndrome, thrombophlebitis migrans, embolism or thrombosis of vena cava (inferior), embolism or thrombosis of renal vein, or mesenteric thrombosis: ICD-9: 452, 453.0, 453.1, 453.2, 453.3, 557.0; ICD-10: I81, I82.0, I82.1, I82.2, I82.3, K55.0H). Additional diagnoses were ITP (ICD-9: 287.31; ICD-10: D69.3), HUS (ICD-9: 283.11; ICD-10: D59.3), and HIT (ICD-9-CM 289.84; ICD-10: D75.82). VTE incidence was first estimated using ICD codes alone and second requiring that an anticoagulant be filled within 30 days (excluding “flush” formulations of heparins) or disenrollment within 30 days of the diagnosis.

Incidence was calculated as the frequency of each diagnosis divided by disease-free person-time across a five-year period. Quarterly incidence was calculated to assess variation and trends across time; monotonic trends were examined using an ordinal correlation coefficient across the 20 quarterly estimates. All enrollees between 26 and 65 years of age with a minimum of 12 consecutive months enrollment were followed from January 1, 2014 to December 31, 2019. Rates were calculated by sex and ten-year age categories: 26-35, 36-45, 46-55, and 56-65. A 12-month event-free lookback period was required for each outcome, before follow-up began. Denominators contained time each enrollee contributed during follow-up (whether or not they used medical services), censored at time of diagnosis, turning 65, or disenrollment. Because Medicare begins at age 65, we limited follow-up to enrollees under 65. There was no patient involvement in this research.

Patient/Public Involvement: None.

## Results

The five-year sample included 13,249,229 people; 6,555,595 were women and 6,693,634 were men, with approximately 3 million people in each of the four age categories (detail in Appendix). Individuals with missing sex (0.01%) or age (0.00007%) information were excluded. For enrollees 25-65 years of age, incidence of a venous thrombotic event (VTE, CVT, or other major venous thrombosis) was 247.89 per 100,000 person-years (95% CI: 245.96, 249.84; 62,598 cases with 25,252,020 person-years). Rates increased with age and were higher for women than for men until age 55, with larger sex differences at younger ages (for all outcomes, see Table for rates per 100,000 for subgroups and overall; see online Appendix for number of cases, person-years, and incidence).

**TABLE:**
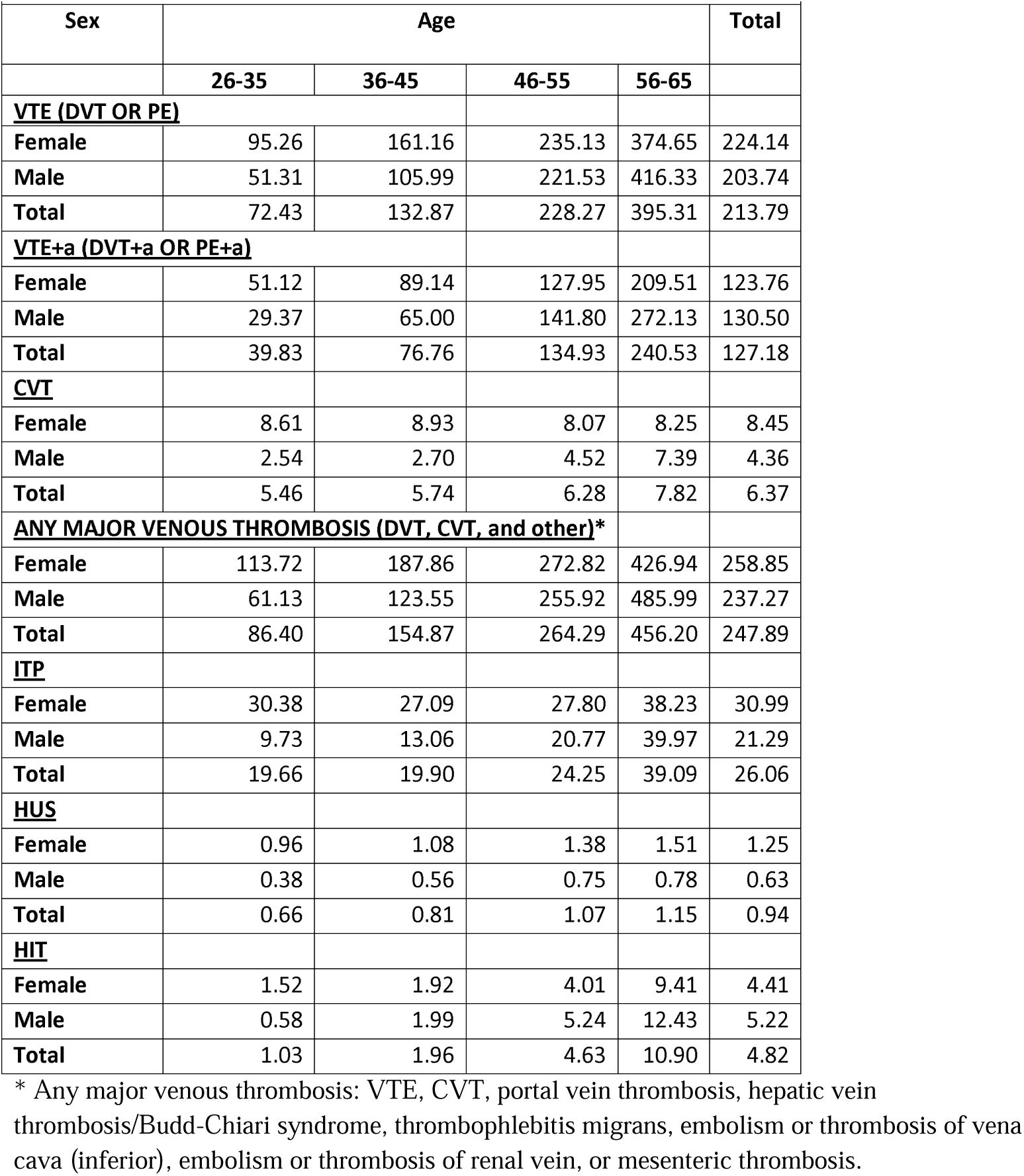
INCIDENCE BY AGE AND SEX PER 100,000 PERSON-YEARS.

| TABLE: INCIDENCE<br>BY AGE AND SEX PER 100,000 PERSON-YEARS |  |  |  |  |  |
| --- | --- | --- | --- | --- | --- |
| Sex | Age |  |  |  | Total |
|  | 26-35 | 36-45 | 46-55 | 56-65 |  |
| **<u>VTE (DVT OR PE)</u>** |  |  |  |  |  |
| Female | 95.26 | 161.16 | 235.13 | 374.65 | 224.14 |
| Male | 51.31 | 105.99 | 221.53 | 416.33 | 203.74 |
| Total | 72.43 | 132.87 | 228.27 | 395.31 | 213.79 |
| **<u>VTE+a (DVT+a OR PE+a)</u>** |  |  |  |  |  |
| Female | 51.12 | 89.14 | 127.95 | 209.51 | 123.76 |
| Male | 29.37 | 65.00 | 141.80 | 272.13 | 130.50 |
| Total | 39.83 | 76.76 | 134.93 | 240.53 | 127.18 |
| **<u>CVT</u>** |  |  |  |  |  |
| Female | 8.61 | 8.93 | 8.07 | 8.25 | 8.45 |
| Male | 2.54 | 2.70 | 4.52 | 7.39 | 4.36 |
| Total | 5.46 | 5.74 | 6.28 | 7.82 | 6.37 |
| **<u>ANY MAJOR VENOUS THROMBOSIS (DVT, CVT, and other)\*</u>** |  |  |  |  |  |
| Female | 113.72 | 187.86 | 272.82 | 426.94 | 258.85 |
| Male | 61.13 | 123.55 | 255.92 | 485.99 | 237.27 |
| Total | 86.40 | 154.87 | 264.29 | 456.20 | 247.89 |
| **<u>ITP</u>** |  |  |  |  |  |
| Female | 30.38 | 27.09 | 27.80 | 38.23 | 30.99 |
| Male | 9.73 | 13.06 | 20.77 | 39.97 | 21.29 |
| Total | 19.66 | 19.90 | 24.25 | 39.09 | 26.06 |
| **<u>HUS</u>** |  |  |  |  |  |
| Female | 0.96 | 1.08 | 1.38 | 1.51 | 1.25 |
| Male | 0.38 | 0.56 | 0.75 | 0.78 | 0.63 |
| Total | 0.66 | 0.81 | 1.07 | 1.15 | 0.94 |
| **<u>HIT</u>** |  |  |  |  |  |
| Female | 1.52 | 1.92 | 4.01 | 9.41 | 4.41 |
| Male | 0.58 | 1.99 | 5.24 | 12.43 | 5.22 |
| Total | 1.03 | 1.96 | 4.63 | 10.90 | 4.82 |

\* Any major venous thrombosis: VTE, CVT, portal vein thrombosis, hepatic vein thrombosis/Budd-Chiari syndrome, thrombophlebitis migrans, embolism or thrombosis of vena cava (inferior), embolism or thrombosis of renal vein, or mesenteric thrombosis.

Incidence of VTE was 213.79 per 100,000 person-years (95% CI: 211.99, 215.59). Incidence of VTE, DVT without PE (47.73; 95% CI: 46.88, 48.58), and PE with or without DVT (171.18; 95% CI: 169.58, 172.80) had similar sex by age patterns with higher rates at older ages and higher rates for women younger than 55 (Appendix). With a more restrictive estimate of VTE (requiring anticoagulation), the incidence of VTE (VTE+a, Table) was 127.18 (95% CI: 125.80, 128.58); 16.32 for DVT alone (95% CI: 15.88, 16.82) and 113.35 for PE with or without DVT (95% CI: 112.05, 114.67). With the restricted definition, rates were higher for women younger than 46.

Quarterly incidence estimates with the restricted definition for DVT, PE, and VTE indicated that DVT decreased significantly across the five-year period (rho = -0.94 women’s total rates, n=20, p<0.0001; rho = -0.98 men’s total rates, n=20, p<0.0001; Figure 1 top panel), while quarterly PE estimates increased significantly (rho = +0.90 women’s total rates, n=20, p<0.0001; rho = +0.66 men’s total rates, n=20, p<0.002; Figure 1 middle panel). The combined VTE quarterly rates showed a significant increase across five years for women (rho = +0.77 total rates, n=20, p<0.0001) and a small, non-significant increase for men (rho = +0.17 total rates, n=20, p<0.47; Figure 1 bottom panel, numeric detail in Appendix). Rates did not appear to be affected by the change from ICD-9 to ICD-10 codes in 2015.

**FIGURE 1:**
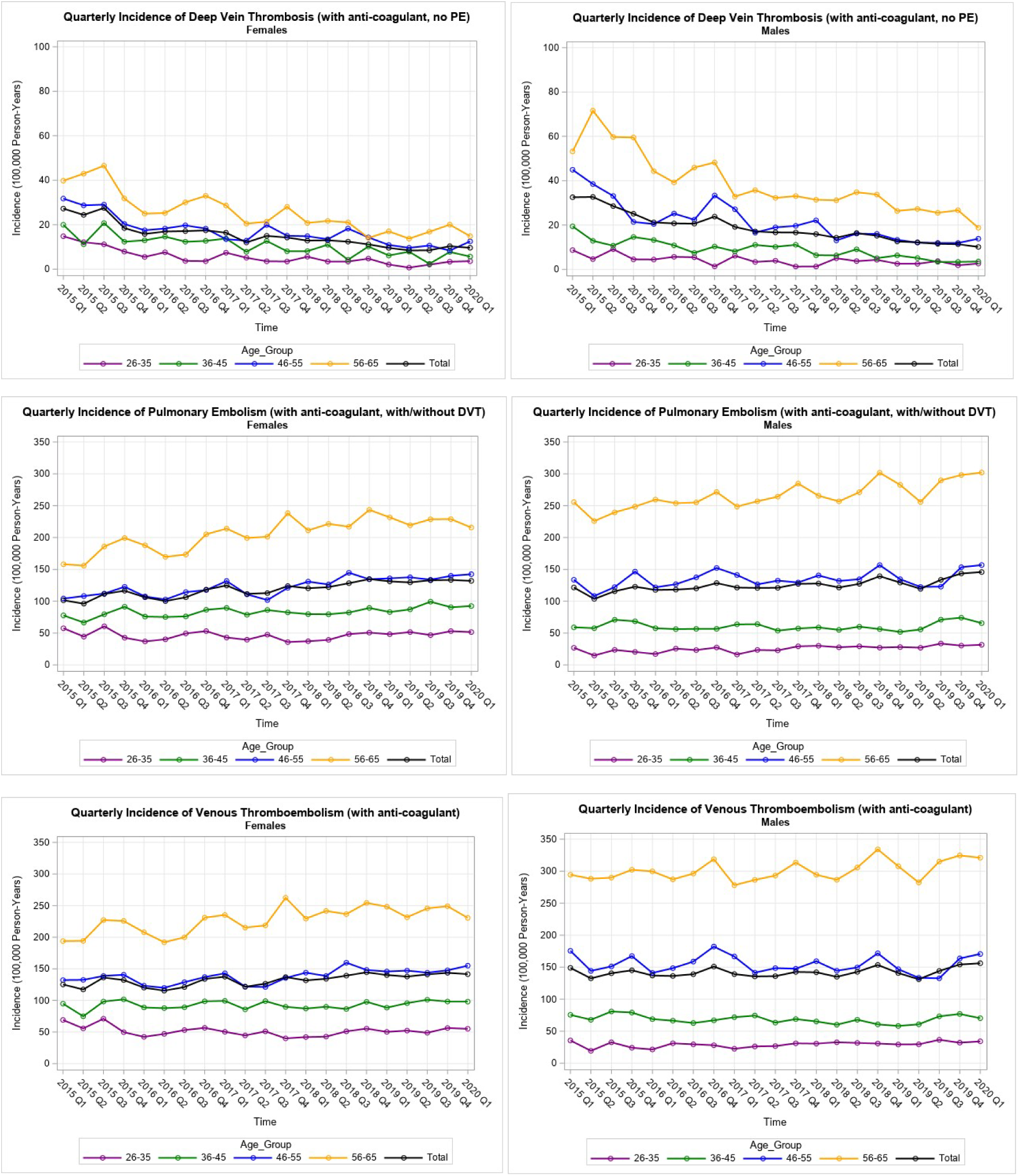
INCIDENCE OF DVT, PE, AND VTE BY SEX AND TIME.

Five-year incidence per 100,000 person-years was 6.37 for CVT (95% CI: 6.07, 6.69), increasing significantly across time for women (rho = +0.66 total rates, n=20, p<0.002) and men (rho = +0.58 total rates, n=20, p<0.01; Figure 2). Five-year incidence for ITP was 26.06 (95% CI: 25.44, 26.78), 4.82 for HIT (95% CI: 4.56, 5.10), and 0.94 for HUS (95% CI: 0.84, 1.06). Incidence was higher in women for all ages for CVT and HUS. Rates for the co-occurrence of CVT with other diagnoses (within 14 days) was 0.04 with ITP (95% CI: 0.02, 0.06), 0.06 with HIT (95% CI:0.03, 0.09), and 0.09 with either ITP or HIT (95% CI: 0.06, 0.13). For the co-occurrence of CVT with other events (Appendix), incidence was higher for women under 55, but was higher for women of all ages for CVT with ITP or HIT.

**FIGURE 2:**
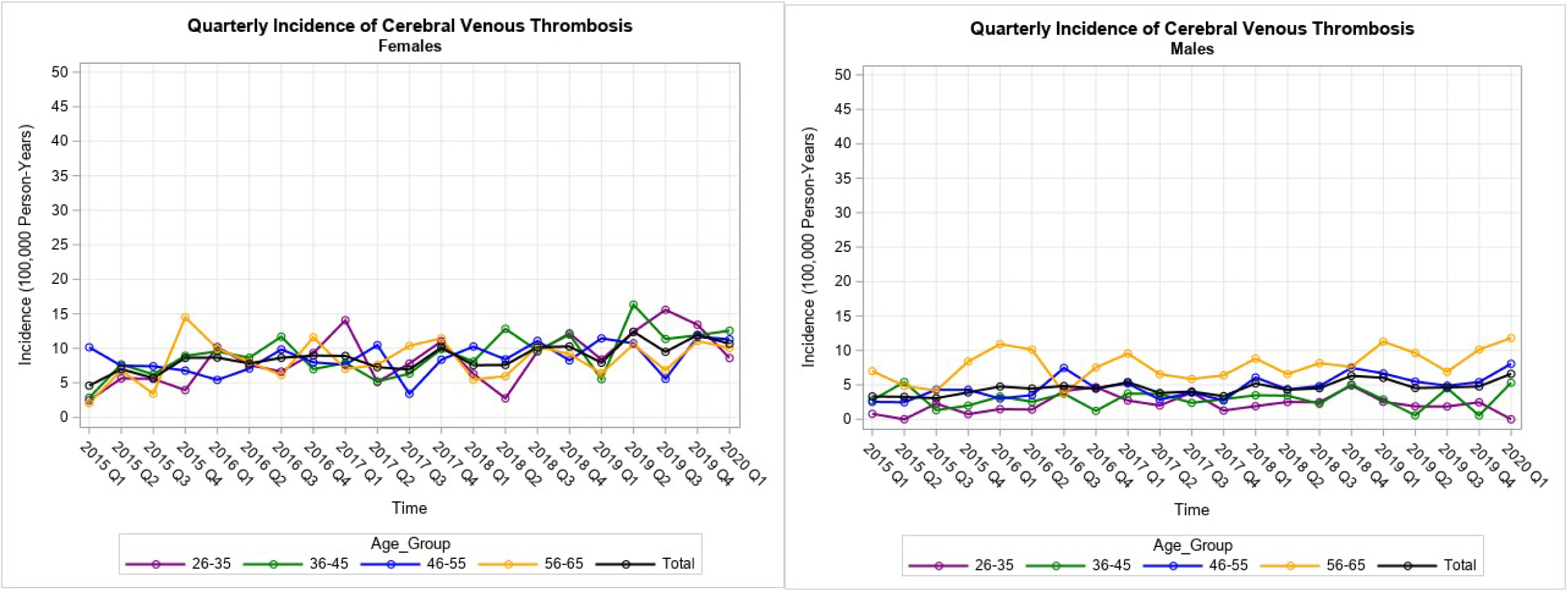
INCIDENCE OF CVT BY SEX AND TIME.

Although it was not part of our trend analysis, rates are shown for the first quarter of 2020 (Figures 1 and 2, and Appendix), because there has been speculation whether the SARS2 coronavirus was circulating in winter 2020 and whether it might elevate rates. Rates for the first quarter of 2020 were not consistently higher than the previous four quarters. Of the eight subgroups, total incidence for PE (and the resulting VTE rate) and CVT were slightly elevated for men.

## Discussion

This is the first report to provide comprehensive age and sex-stratified incidence estimates for venous thromboembolic and other rare events of interest in the United States, although the cohort is limited to insured enrollees and does not include those uninsured or with other insurance carriers. Estimates cover the most recent period before the COVID-19 pandemic. Rates increased with older age for most outcomes and tended to be higher for women under 55. Incidence for CVT, HUS, and CVT with ITP or HIT was higher for women at all ages.

Incidence for VTE ranged from 127 per 100,000 person-years when an anticoagulant was required to 214 when using only ICD codes. The higher estimate includes cases, as well as suspected cases, and likely overestimates the true rate. It is, however, the method used in other studies.[5] The lower estimate may underestimate the true incidence as anyone receiving – but not filling – their anticoagulant prescription would be excluded, which may occur as often as in 40% of cases.[8] Thus, results may be limited by use of administrative-level data and without detailed patient records. However, a recent cohort study of Kaiser-Permanente patients (insured, with complete patient records) compared COVID-19 positive with COVID-19 negative patients and estimated the incidence per 100,000 of VTE in COVID-19 negative patients (n∼200,000) as 160 for outpatients and 300 for hospital-associated diagnoses (inpatient or post-hospital).[9] Thus, these estimates, which are also higher than European rates, suggest our estimates may provide upper and lower bounds on true VTE incidence in the United States. Population differences in rates are notable, as confidence intervals between studies often do not overlap. In the past decade, VTE incidence per 100,000 person-years ranged from 150 in the UK[6] to 200 in Spain[6] for adults of all ages (Figure 3; study details in Appendix).[4–6,10–16] A recent report from Denmark[5] estimated incidence (2010-2018) for VTE at 170 and 176 for a broader category of venous thromboses among those 18-99 years of age in a population base of five million. For those 18-64 years of age, incidence was 91 for VTE and 95 for the more inclusive category. In this study, rates for similar age and time ranges were twice as high (214 for VTE, 248 including other major venous thrombotic events).

**Figure 3.**
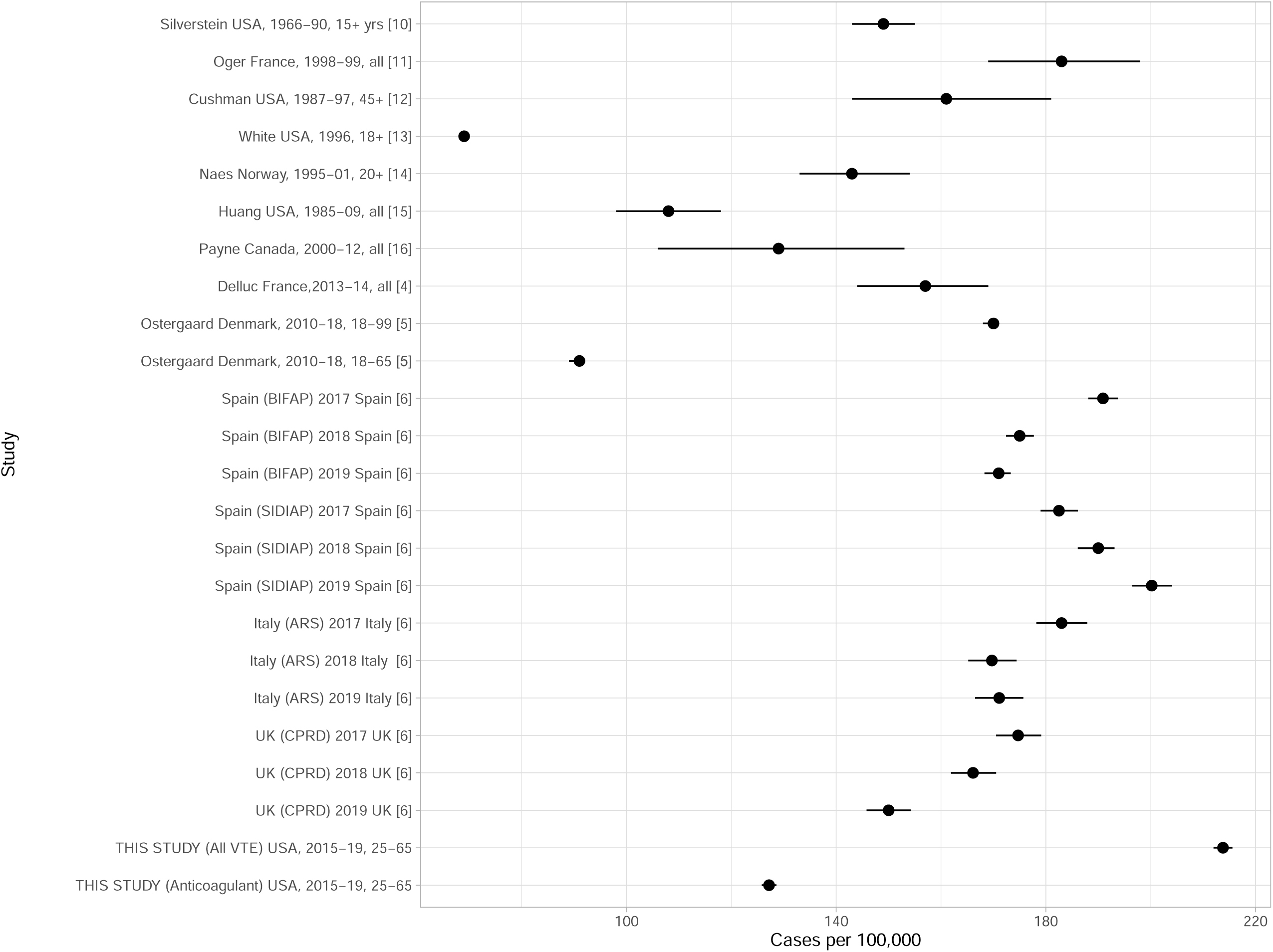
VTE Incidence across Countries and Time per 100,000

CVT estimates may appear more consistent across populations, but also may have non-overlapping confidence intervals. In the past decade, CVT incidence per 100,000 ranged from 0.12 in the UK[6] to 2.00 in New York/Florida in the US[17] (Figure 4;[6,17–19] study details in Appendix). In this analysis, CVT incidence (6.37) was several times higher than reported in European countries and three times higher than New York/Florida. In contrast, the ITP rate in this study (26.06) fell within the range of 2019 European rates from 20.12 in the UK to 95.71 in Spain.[6] The previous US study[17] used hospital discharge data and estimated state populations to estimate CVT incidence, while this study used diagnoses occurring in any setting in a well-defined cohort; but both studies found increased incidence over time. Validation studies indicate high positive predictive value (87-100%) for the CVT ICD-9 and ICD-10 codes used in this study, with the exception of 437.6 (33%),[20,21] but additionally indicate some cases were outpatient diagnoses.[20]

**Figure 4.**
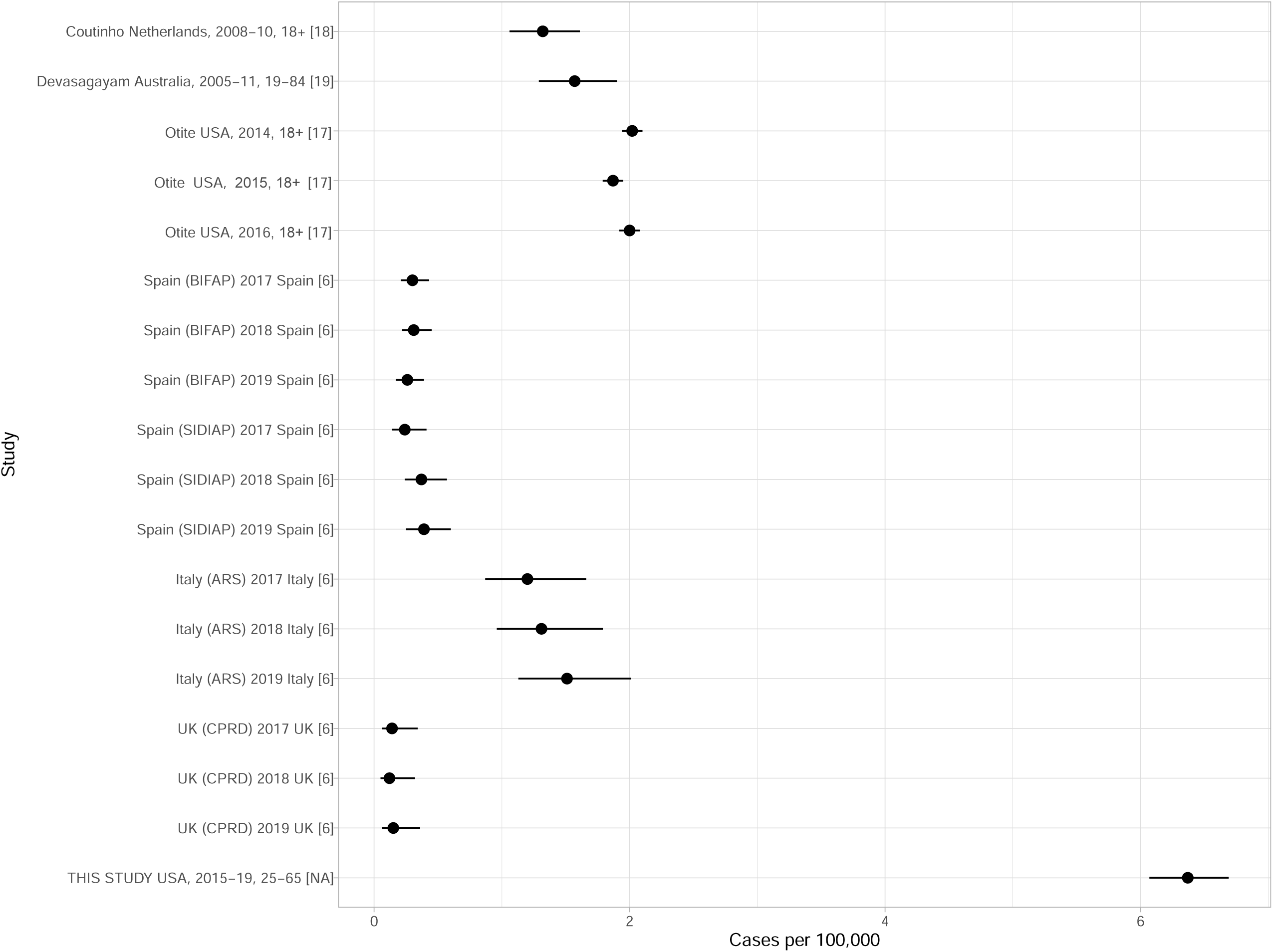
CVT Incidence across Countries and Time per 100,000

Trends though time indicated that in the past five years, US rates for DVT decreased significantly and PE rates increased significantly for both sexes, but the aggregate VTE rates showed an increase only for women. There has been speculation that the SARS2 coronavirus may have elevated thrombotic events as it began circulating in winter 2020. Although this is outside of our original protocol and one data point is not sufficient to discern if there has been a change in trend, we presented the rates for the first quarter in 2020. The first quarter rates for 2020 appeared elevated only for men for PE (and VTE) and CVT, but these rates appear within a series of rates for PE and CVT that have been increasing across time. More data would be necessary to discern whether the rate of increase (slope) was greater in 2020.

Inconsistencies across studies may be due to methodological differences between studies and to differences in underlying population characteristics (age, smoking, obesity, and comorbidities), lack of standardization, differences in included ICD codes, or detection bias. To be valid, a comparison of population-based rates must at a minimum be similar in several critical areas such as time periods, and be standardized to a reference population on those characteristics for which data are available such as age and sex. Use of a commercially insured cohort to estimate incidence has limited generalizability to the entire US population (unlike studies using national health care system data). People in the cohort tend to be employed and the uninsured are excluded. Exclusion of the uninsured would likely result in slightly higher estimates of non-life-threatening health service usage. The OPTUM cohort has greater representation of those residing in the southern states, which could increase inclusion of those who smoke and/or are obese. Also, estimates based on administrative data not linked to patient records could overestimate rates by including suspected/probable cases among the definite cases. However, VTE estimates appeared to be similar to those from an insured population with access to complete patient records[9] and CVT codes had good positive predictive value.[20,21] Administrative data also are limited in the availability of reliable sociodemographic information, including obesity and smoking. Finally, although we excluded any prior CVT, our CVT case count might be slightly overestimated by not excluding those who had an IVC filter placed within the preceding year. We also did not report the co-morbidities of the cases. Future research may attempt meta-analyses of studies with similar methodologies, as was done in Canada.[16]

This is the first comprehensive US report on incidence of these conditions, since advances have been made in the past decade in thromboprophylaxis and detection. Although generalizations to the US population from this dataset may be limited, even with such a large dataset, because it is not representative of the US population, there are few longitudinal US databases with the level of necessary detail for those younger than 65. Thus, the Optum database is one of the few options for estimating rarer health events in the US population and has been used for detection of vaccine adverse events.[22] Furthermore, this report contains some of the first population-based rates for the incidence of rare events associated with thrombocytopenia. Rates may be useful when determining excess morbidity linked to COVID-19 and more generally for vaccine adverse event surveillance.[22–24]

## Data Availability

All available data appear in either the figures or Appendix of this study.

## Data Sharing

No further data are available. Data are proprietary of OPTUM Health Systems. However, requests for re-analyses will be considered.

## Ethics

This study was approved by the University of Texas Medical Branch at Galveston’s Institutional Review Board (IRB #20-0313).

## Author Contributorship

Weller acquired grant funding for this project, drafted the manuscript, and was responsible for the design and implementation of the project. Baillargeon and Wilkinson contributed to the design and implementation of the project. Porterfield provided clinical expertise on the project with regard to diagnoses, diagnostic categories, and interpretation. Davis participated in the systematic review of previous incidence studies, assisted with selection of diagnostic codes, and conducted statistical analyses. Chen was the primary data coder and was responsible for data integrity and calculation of incidence estimates. All authors contributed to writing and editing of the manuscript and approved the final draft.

