## Supplement for "INCIDENCE OF VENOUS THROMBOTIC EVENTS AND EVENTS OF SPECIAL INTEREST IN A RETROSPECTIVE COHORT OF COMMERCIALLY-INSURED US PATIENTS"

**ONLINE APPENDIX:**

**CASE COUNTS AND PERSON-TIME EXPOSURES FOR EACH OUTCOME BY AGE AND SEX STRATA**

**Page	Contents**

1 **TABLE OF CONTENTS**
2 **FIVE YEAR INCIDENCE RATES (AND SAMPLE SIZES)**
  a. DVT Incidence
  b. PE Incidence
  c. VTE Incidence
  d. DVT Incidence with anticoagulant
  e. PE Incidence with anticoagulant
  f. VTE Incidence with anticoagulant
  g. CVT Incidence
  h. Any Major Venous Thrombosis Incidence (DVT, CVT, and other)
  i. ITP Incidence
  j. CVT and ITP incidence of co-occurrence
  k. HUS Incidence
  l. HIT Incidence
  m. CVT and HIT incidence of co-occurrence
  n. CVT and (ITP or HIT) incidence of co-occurrence
16 **QUARTERLY INCIDENCE RATES (by type of thrombosis, year, sex, and age)**

**48	Incidence SUMMARY with FIRST Quarter 2020**

**49	SYSTEMATIC REVIEW TABLES FOR INCIDENCE ESTIMATES ACROSS STUDIES**

a. Summary of studies for VTE incidence
b. VTE references
c. Table of studies with CVT incidence
d. CVT references

**FIVE YEAR INCIDENCE RATES: SAMPLE SIZES BY SEX AND AGE**

| **Sex** | **Age** | | | | **Total** |
| --- | --- | --- | --- | --- | --- |
|  | **26-35** | **36-45** | **46-55** | **56-65** |  |
| **F** | 1773785 | 1606766 | 1661974 | 1513070 | 6555595 |
| **M** | 1856141 | 1679084 | 1677182 | 1481227 | 6693634 |
| **Total** | 3629926 | 3285850 | 3339156 | 2994297 | 1.325E7 |

Anyone missing sex (0.01%) or age (6.65411E-05%) were excluded.

| **DVT** | **Age Group** | **DVT1 Cases** | **Person-Years** | **Incidence (Cases per 100,000 Person-Year)** |
| --- | --- | --- | --- | --- |
| **Female** | **26-35** | 541 | 2,693,323 | 20.09 |
|  | **36-45** | 1,161 | 3,063,674 | 37.90 |
|  | **46-55** | 1,900 | 3,388,315 | 56.08 |
|  | **56-65** | 2,734 | 3,360,666 | 81.35 |
|  | **Total** | 6,336 | 12,505,977 | 50.66 |
| **Male** | **26-35** | 355 | 2,908,750 | 12.20 |
|  | **36-45** | 832 | 3,221,362 | 25.83 |
|  | **46-55** | 1,755 | 3,448,322 | 50.89 |
|  | **56-65** | 2,840 | 3,305,545 | 85.92 |
|  | **Total** | 5,782 | 12,883,979 | 44.88 |
| **Total** | **26-35** | 896 | 5,602,073 | 15.99 |
|  | **36-45** | 1,993 | 6,285,036 | 31.71 |
|  | **46-55** | 3,655 | 6,836,636 | 53.46 |
|  | **56-65** | 5,574 | 6,666,211 | 83.62 |
|  | **Total** | 12,118 | 25,389,957 | 47.73 |
| CASE is acute deep vein thrombosis (without PE) | | | | |

| **PE** | **Age Group** | **PE1 Cases** | **Person-Years** | **Incidence (Cases per 100,000 Person-Year)** |
| --- | --- | --- | --- | --- |
| **Female** | **26-35** | 2,046 | 2,689,876 | 76.06 |
|  | **36-45** | 3,860 | 3,057,222 | 126.26 |
|  | **46-55** | 6,221 | 3,377,408 | 184.19 |
|  | **56-65** | 10,093 | 3,342,999 | 301.91 |
|  | **Total** | 22,220 | 12,467,506 | 178.22 |
| **Male** | **26-35** | 1,161 | 2,907,075 | 39.94 |
|  | **36-45** | 2,663 | 3,217,161 | 82.77 |
|  | **46-55** | 6,063 | 3,437,737 | 176.37 |
|  | **56-65** | 11,226 | 3,284,586 | 341.78 |
|  | **Total** | 21,113 | 12,846,560 | 164.35 |
| **Total** | **26-35** | 3,207 | 5,596,952 | 57.30 |
|  | **36-45** | 6,523 | 6,274,384 | 103.96 |
|  | **46-55** | 12,284 | 6,815,145 | 180.25 |
|  | **56-65** | 21,319 | 6,627,586 | 321.67 |
|  | **Total** | 43,333 | 25,314,066 | 171.18 |
| CASE is acute pulmonary embolism  (with or without DVT) | | | | |

| **VTE** | **Age Group** | **VTE1 Cases** | **Person-Years** | **Incidence (Cases per 100,000 Person-Year)** |
| --- | --- | --- | --- | --- |
| **Female** | **26-35** | 2,561 | 2,688,417 | 95.26 |
|  | **36-45** | 4,921 | 3,053,471 | 161.16 |
|  | **46-55** | 7,926 | 3,370,917 | 235.13 |
|  | **56-65** | 12,489 | 3,333,493 | 374.65 |
|  | **Total** | 27,897 | 12,446,299 | 224.14 |
| **Male** | **26-35** | 1,491 | 2,906,144 | 51.31 |
|  | **36-45** | 3,407 | 3,214,526 | 105.99 |
|  | **46-55** | 7,603 | 3,431,977 | 221.53 |
|  | **56-65** | 13,633 | 3,274,559 | 416.33 |
|  | **Total** | 26,134 | 12,827,206 | 203.74 |
| **Total** | **26-35** | 4,052 | 5,594,561 | 72.43 |
|  | **36-45** | 8,328 | 6,267,998 | 132.87 |
|  | **46-55** | 15,529 | 6,802,894 | 228.27 |
|  | **56-65** | 26,122 | 6,608,052 | 395.31 |
|  | **Total** | 54,031 | 25,273,505 | 213.79 |
| CASE is DVT or PE | |  |  |  |

| **DVT & anti-coagulant** | **Age Group** | **DVT2 Cases** | **Person-Years** | **Incidence (Cases per 100,000 Person-Year)** |
| --- | --- | --- | --- | --- |
| **Female** | **26-35** | 147 | 2,694,422 | 5.46 |
|  | **36-45** | 324 | 3,066,547 | 10.57 |
|  | **46-55** | 556 | 3,393,038 | 16.39 |
|  | **56-65** | 813 | 3,367,677 | 24.14 |
|  | **Total** | 1,840 | 12,521,684 | 14.69 |
| **Male** | **26-35** | 123 | 2,909,358 | 4.23 |
|  | **36-45** | 286 | 3,223,177 | 8.87 |
|  | **46-55** | 705 | 3,451,828 | 20.42 |
|  | **56-65** | 1,194 | 3,311,639 | 36.05 |
|  | **Total** | 2,308 | 12,896,003 | 17.90 |
| **Total** | **26-35** | 270 | 5,603,781 | 4.82 |
|  | **36-45** | 610 | 6,289,724 | 9.70 |
|  | **46-55** | 1,261 | 6,844,866 | 18.42 |
|  | **56-65** | 2,007 | 6,679,316 | 30.05 |
|  | **Total** | 4,148 | 25,417,687 | 16.32 |
| CASE is acute deep vein thrombosis (without PE) AND (anticoagulant Rx within 30d OR disenrollment in 30d) | | | | |

| **PE & anti-coagulant** | **Age Group** | **PE2 Cases** | **Person-Years** | **Incidence (Cases per 100,000 Person-Year)** |
| --- | --- | --- | --- | --- |
| **Female** | **26-35** | 1,242 | 2,692,042 | 46.14 |
|  | **36-45** | 2,449 | 3,061,395 | 80.00 |
|  | **46-55** | 3,859 | 3,384,485 | 114.02 |
|  | **56-65** | 6,358 | 3,354,104 | 189.56 |
|  | **Total** | 13,908 | 12,492,026 | 111.34 |
| **Male** | **26-35** | 742 | 2,908,127 | 25.51 |
|  | **36-45** | 1,844 | 3,219,535 | 57.28 |
|  | **46-55** | 4,274 | 3,443,294 | 124.13 |
|  | **56-65** | 7,976 | 3,295,074 | 242.06 |
|  | **Total** | 14,836 | 12,866,030 | 115.31 |
| **Total** | **26-35** | 1,984 | 5,600,169 | 35.43 |
|  | **36-45** | 4,293 | 6,280,931 | 68.35 |
|  | **46-55** | 8,133 | 6,827,779 | 119.12 |
|  | **56-65** | 14,334 | 6,649,178 | 215.58 |
|  | **Total** | 28,744 | 25,358,056 | 113.35 |
| CASE is PE (with or without DVT) AND (anticoagulant Rx within 30d OR disenrollment in 30d) | | | | |

| **VTE & anti-coagulant** | **Age Group** | **VTE2 Cases** | **Person-Years** | **Incidence (Cases per 100,000 Person-Year)** |
| --- | --- | --- | --- | --- |
| **Female** | **26-35** | 1,376 | 2,691,654 | 51.12 |
|  | **36-45** | 2,728 | 3,060,437 | 89.14 |
|  | **46-55** | 4,328 | 3,382,522 | 127.95 |
|  | **56-65** | 7,021 | 3,351,228 | 209.51 |
|  | **Total** | 15,453 | 12,485,841 | 123.76 |
| **Male** | **26-35** | 854 | 2,907,779 | 29.37 |
|  | **36-45** | 2,092 | 3,218,617 | 65.00 |
|  | **46-55** | 4,879 | 3,440,821 | 141.8 |
|  | **56-65** | 8,955 | 3,290,667 | 272.13 |
|  | **Total** | 16,780 | 12,857,884 | 130.5 |
| **Total** | **26-35** | 2,230 | 5,599,433 | 39.83 |
|  | **36-45** | 4,820 | 6,279,054 | 76.76 |
|  | **46-55** | 9,207 | 6,823,343 | 134.93 |
|  | **56-65** | 15,976 | 6,641,895 | 240.53 |
|  | **Total** | 32,233 | 25,343,725 | 127.18 |
| CASE is DVT and/or PE AND (anticoagulant Rx within 30d OR disenrollment in 30d) | | | | |

| **CVT** | **Age Group** | **CVT Cases** | **Person-Years** | **Incidence (Cases per 100,000 Person-Year)** |
| --- | --- | --- | --- | --- |
| **Female** | **26-35** | 232 | 2,694,450 | 8.61 |
|  | **36-45** | 274 | 3,067,010 | 8.93 |
|  | **46-55** | 274 | 3,394,583 | 8.07 |
|  | **56-65** | 278 | 3,370,274 | 8.25 |
|  | **Total** | 1,058 | 12,526,317 | 8.45 |
| **Male** | **26-35** | 74 | 2,909,606 | 2.54 |
|  | **36-45** | 87 | 3,223,999 | 2.70 |
|  | **46-55** | 156 | 3,454,185 | 4.52 |
|  | **56-65** | 245 | 3,316,001 | 7.39 |
|  | **Total** | 562 | 12,903,791 | 4.36 |
| **Total** | **26-35** | 306 | 5,604,057 | 5.46 |
|  | **36-45** | 361 | 6,291,008 | 5.74 |
|  | **46-55** | 430 | 6,848,768 | 6.28 |
|  | **56-65** | 523 | 6,686,274 | 7.82 |
|  | **Total** | 1,620 | 25,430,108 | 6.37 |
| CASE is CVT with or without other diagnoses | | | |  |

| **ANY Major Venous Thrombosis** | **Age Group** | **ANY thrombosis Cases** | **Person-Years** | **Incidence (Cases per 100,000 Person-Year)** |
| --- | --- | --- | --- | --- |
| **Female** | **26-35** | 3,056 | 2,687,306 | 113.72 |
|  | **36-45** | 5,732 | 3,051,277 | 187.86 |
|  | **46-55** | 9,187 | 3,367,448 | 272.82 |
|  | **56-65** | 14,212 | 3,328,824 | 426.94 |
|  | **Total** | 32,187 | 12,434,854 | 258.85 |
| **Male** | **26-35** | 1,776 | 2,905,443 | 61.13 |
|  | **36-45** | 3,970 | 3,213,162 | 123.55 |
|  | **46-55** | 8,776 | 3,429,143 | 255.92 |
|  | **56-65** | 15,889 | 3,269,418 | 485.99 |
|  | **Total** | 30,411 | 12,817,166 | 237.27 |
| **Total** | **26-35** | 4,832 | 5,592,749 | 86.40 |
|  | **36-45** | 9,702 | 6,264,439 | 154.87 |
|  | **46-55** | 17,963 | 6,796,591 | 264.29 |
|  | **56-65** | 30,101 | 6,598,242 | 456.20 |
|  | **Total** | 62,598 | 25,252,020 | 247.89 |
| Any of the above (DVT, PE, CVT or Any of the following: ) | | | |  |
| Portal vein thrombosis | | |  |  |
| Hepatic vein thrombosis/Budd-Chiari syndrome | | |  |  |
| Thrombophlebitis migrans | | |  |  |
| Embolism or thrombosis of vena cava (inferior) | | |  |  |
| Embolism or thrombosis of renal vein | | |  |  |
| Mesenteric thrombosis | | |  |  |

| **ITP** | **Age Group** | **ITP Cases** | **Person-Years** | **Incidence (Cases per 100,000 Person-Year)** |
| --- | --- | --- | --- | --- |
| **Female** | **26-35** | 818 | 2,692,331 | 30.38 |
|  | **36-45** | 830 | 3,064,415 | 27.09 |
|  | **46-55** | 943 | 3,391,788 | 27.80 |
|  | **56-65** | 1,287 | 3,366,206 | 38.23 |
|  | **Total** | 3,878 | 12,514,741 | 30.99 |
| **Male** | **26-35** | 283 | 2,908,880 | 9.73 |
|  | **36-45** | 421 | 3,222,809 | 13.06 |
|  | **46-55** | 717 | 3,452,164 | 20.77 |
|  | **56-65** | 1,324 | 3,312,432 | 39.97 |
|  | **Total** | 2,745 | 12,896,285 | 21.29 |
| **Total** | **26-35** | 1,101 | 5,601,211 | 19.66 |
|  | **36-45** | 1,251 | 6,287,224 | 19.90 |
|  | **46-55** | 1,660 | 6,843,952 | 24.25 |
|  | **56-65** | 2,611 | 6,678,638 | 39.09 |
|  | **Total** | 6,623 | 25,411,025 | 26.06 |
| Case is Immune thrombocytopenic purpura | | | |  |

| **CVT+ITP** | **Age Group** | **CVT AND ITP Cases** | **Person-Years** | **Incidence (Cases per 1,000,000 Person-Year)** |
| --- | --- | --- | --- | --- |
| **Female** | **26-35** | 3 | 2,694,835 | 1.11 |
|  | **36-45** | 2 | 3,067,560 | 0.65 |
|  | **46-55** | 2 | 3,395,209 | 0.59 |
|  | **56-65** | 0 | 3,370,847 | 0.00 |
|  | **Total** | 7 | 12,528,452 | 0.56 |
| **Male** | **26-35** | 0 | 2,909,727 | 0.00 |
|  | **36-45** | 1 | 3,224,171 | 0.31 |
|  | **46-55** | 1 | 3,454,477 | 0.29 |
|  | **56-65** | 0 | 3,316,471 | 0.00 |
|  | **Total** | 2 | 12,904,846 | 0.15 |
| **Total** | **26-35** | 3 | 5,604,563 | 0.54 |
|  | **36-45** | 3 | 6,291,731 | 0.48 |
|  | **46-55** | 3 | 6,849,686 | 0.44 |
|  | **56-65** | 0 | 6,687,318 | 0.00 |
|  | **Total** | 9 | 25,433,298 | 0.35 |
| CASE is CVT AND ITP (as above, if diagnoses are within 14 days of each other) | | |  |  |

| **HUS** | **Age Group** | **HUS Cases** | **Person-Years** | **Incidence (Cases per 1,000,000 Person-Year)** |
| --- | --- | --- | --- | --- |
| **Female** | **26-35** | 26 | 2,694,772 | 9.65 |
|  | **36-45** | 33 | 3,067,490 | 10.76 |
|  | **46-55** | 47 | 3,395,111 | 13.84 |
|  | **56-65** | 51 | 3,370,714 | 15.13 |
|  | **Total** | 157 | 12,528,087 | 12.53 |
| **Male** | **26-35** | 11 | 2,909,708 | 3.78 |
|  | **36-45** | 18 | 3,224,124 | 5.58 |
|  | **46-55** | 26 | 3,454,429 | 7.53 |
|  | **56-65** | 26 | 3,316,407 | 7.84 |
|  | **Total** | 81 | 12,904,668 | 6.28 |
| **Total** | **26-35** | 37 | 5,604,480 | 6.60 |
|  | **36-45** | 51 | 6,291,614 | 8.11 |
|  | **46-55** | 73 | 6,849,540 | 10.66 |
|  | **56-65** | 77 | 6,687,121 | 11.51 |
|  | **Total** | 238 | 25,432,755 | 9.36 |
| Case is Hemolytic-uremic syndrome | | |  |  |

| **HIT** | **Age Group** | **HIT Cases** | **Person-Years** | **Incidence (Cases per 100,000 Person-Year)** |
| --- | --- | --- | --- | --- |
| **Female** | **26-35** | 41 | 2,694,757 | 1.52 |
|  | **36-45** | 59 | 3,067,445 | 1.92 |
|  | **46-55** | 136 | 3,394,948 | 4.01 |
|  | **56-65** | 317 | 3,370,282 | 9.41 |
|  | **Total** | 553 | 12,527,432 | 4.41 |
| **Male** | **26-35** | 17 | 2,909,692 | 0.58 |
|  | **36-45** | 64 | 3,224,044 | 1.99 |
|  | **46-55** | 181 | 3,454,172 | 5.24 |
|  | **56-65** | 412 | 3,315,697 | 12.43 |
|  | **Total** | 674 | 12,903,605 | 5.22 |
| **Total** | **26-35** | 58 | 5,604,449 | 1.03 |
|  | **36-45** | 123 | 6,291,489 | 1.96 |
|  | **46-55** | 317 | 6,849,120 | 4.63 |
|  | **56-65** | 729 | 6,685,980 | 10.90 |
|  | **Total** | 1,227 | 25,431,037 | 4.82 |
| CASE is HIT | |  |  |  |

| **CVT+HIT** | **Age Group** | **CVT AND HIT Cases** | **Person-Years** | **Incidence (Cases per 1,000,000 Person-Year)** |
| --- | --- | --- | --- | --- |
| **Female** | **26-35** | 1 | 2,694,838 | 0.37 |
|  | **36-45** | 1 | 3,067,566 | 0.33 |
|  | **46-55** | 3 | 3,395,208 | 0.88 |
|  | **56-65** | 6 | 3,370,841 | 1.78 |
|  | **Total** | 11 | 12,528,454 | 0.88 |
| **Male** | **26-35** | 0 | 2,909,727 | 0.00 |
|  | **36-45** | 2 | 3,224,170 | 0.62 |
|  | **46-55** | 0 | 3,454,478 | 0.00 |
|  | **56-65** | 2 | 3,316,468 | 0.60 |
|  | **Total** | 4 | 12,904,843 | 0.31 |
| **Total** | **26-35** | 1 | 5,604,566 | 0.18 |
|  | **36-45** | 3 | 6,291,736 | 0.48 |
|  | **46-55** | 3 | 6,849,686 | 0.44 |
|  | **56-65** | 8 | 6,687,310 | 1.20 |
|  | **Total** | 15 | 25,433,297 | 0.59 |
| CASE is CVT AND HIT (as above, If diagnoses are within 14 days of each other) | | |  |  |

| **CVT +**  **(ITP or HIT)** | **Age Group** | **CVT AND (ITP or HIT) Cases** | **Person-Years** | **Incidence (Cases per 1,000,000 Person-Year)** |
| --- | --- | --- | --- | --- |
| **Female** | **26-35** | 4 | 2,694,835 | 1.48 |
|  | **36-45** | 3 | 3,067,560 | 0.98 |
|  | **46-55** | 5 | 3,395,203 | 1.47 |
|  | **56-65** | 6 | 3,370,841 | 1.78 |
|  | **Total** | 18 | 12,528,439 | 1.44 |
| **Male** | **26-35** | 0 | 2,909,727 | 0.00 |
|  | **36-45** | 2 | 3,224,170 | 0.62 |
|  | **46-55** | 1 | 3,454,477 | 0.29 |
|  | **56-65** | 2 | 3,316,468 | 0.60 |
|  | **Total** | 5 | 12,904,842 | 0.39 |
| **Total** | **26-35** | 4 | 5,604,563 | 0.71 |
|  | **36-45** | 5 | 6,291,730 | 0.79 |
|  | **46-55** | 6 | 6,849,680 | 0.88 |
|  | **56-65** | 8 | 6,687,310 | 1.20 |
|  | **Total** | 23 | 25,433,282 | 0.90 |
| CASE is CVT AND (ITP or HIT) | | |  |  |
| If diagnoses are within 14 days of each other | | |  |  |

**QUARTERLY INCIDENCE RATES**

**QUARTERLY INCIDENCE RATES: DVT with anticoagulant 2015 (Cases per 100,000 Person-Years)**

|  |  | **2015 Q1** | | | **2015 Q2** | | | **2015 Q3** | | | **2015 Q4** | | |
| --- | --- | --- | --- | --- | --- | --- | --- | --- | --- | --- | --- | --- | --- |
|  | **Age Group** | **Cases** | **Person-Years** | **Incidence** | **Cases** | **Person-Years** | **Incidence** | **Cases** | **Person-Years** | **Incidence** | **Cases** | **Person-Years** | **Incidence** |
| **Female** | **26-35** | 18 | 121,688 | 14.79 | 15 | 123,878 | 12.11 | 14 | 125,384 | 11.17 | 10 | 126,592 | 7.9 |
|  | **36-45** | 28 | 140,419 | 19.94 | 16 | 143,125 | 11.18 | 30 | 144,747 | 20.73 | 18 | 145,735 | 12.35 |
|  | **46-55** | 50 | 157,642 | 31.72 | 46 | 160,283 | 28.7 | 47 | 161,819 | 29.04 | 33 | 162,498 | 20.31 |
|  | **56-65** | 58 | 145,882 | 39.76 | 63 | 146,667 | 42.95 | 68 | 146,170 | 46.52 | 46 | 144,888 | 31.75 |
|  | **Total** | 154 | 565,631 | 27.23 | 140 | 573,952 | 24.39 | 159 | 578,119 | 27.5 | 107 | 579,713 | 18.46 |
| **Male** | **26-35** | 11 | 126,587 | 8.69 | 6 | 129,366 | 4.64 | 12 | 131,745 | 9.11 | 6 | 132,695 | 4.52 |
|  | **36-45** | 28 | 144,358 | 19.4 | 19 | 147,527 | 12.88 | 16 | 149,815 | 10.68 | 22 | 150,564 | 14.61 |
|  | **46-55** | 71 | 158,070 | 44.92 | 62 | 161,177 | 38.47 | 54 | 163,109 | 33.11 | 35 | 163,461 | 21.41 |
|  | **56-65** | 76 | 142,848 | 53.2 | 103 | 143,914 | 71.57 | 86 | 144,052 | 59.7 | 85 | 142,887 | 59.49 |
|  | **Total** | 186 | 571,862 | 32.53 | 190 | 581,983 | 32.65 | 168 | 588,721 | 28.54 | 148 | 589,607 | 25.1 |
| **Total** | **26-35** | 29 | 248,275 | 11.68 | 21 | 253,243 | 8.29 | 26 | 257,129 | 10.11 | 16 | 259,287 | 6.17 |
|  | **36-45** | 56 | 284,777 | 19.66 | 35 | 290,652 | 12.04 | 46 | 294,562 | 15.62 | 40 | 296,299 | 13.5 |
|  | **46-55** | 121 | 315,712 | 38.33 | 108 | 321,460 | 33.6 | 101 | 324,928 | 31.08 | 68 | 325,959 | 20.86 |
|  | **56-65** | 134 | 288,730 | 46.41 | 166 | 290,580 | 57.13 | 154 | 290,222 | 53.06 | 131 | 287,775 | 45.52 |
|  | **Total** | 340 | 1,137,494 | 29.89 | 330 | 1,155,935 | 28.55 | 327 | 1,166,841 | 28.02 | 255 | 1,169,320 | 21.81 |

**QUARTERLY INCIDENCE RATES: DVT with anticoagulant 2016 (Cases per 100,000 Person-Years)**

|  |  | **2016 Q1** | | | **2016 Q2** | | | **2016 Q3** | | | **2016 Q4** | | |
| --- | --- | --- | --- | --- | --- | --- | --- | --- | --- | --- | --- | --- | --- |
|  | **Age Group** | **Cases** | **Person-Years** | **Incidence** | **Cases** | **Person-Years** | **Incidence** | **Cases** | **Person-Years** | **Incidence** | **Cases** | **Person-Years** | **Incidence** |
| **Female** | **26-35** | 7 | 127,608 | 5.49 | 10 | 131,901 | 7.58 | 5 | 135,675 | 3.69 | 5 | 139,624 | 3.58 |
|  | **36-45** | 19 | 146,541 | 12.97 | 22 | 150,471 | 14.62 | 19 | 153,913 | 12.34 | 20 | 157,489 | 12.7 |
|  | **46-55** | 29 | 165,896 | 17.48 | 31 | 169,652 | 18.27 | 34 | 172,690 | 19.69 | 32 | 175,575 | 18.23 |
|  | **56-65** | 40 | 160,221 | 24.97 | 41 | 162,455 | 25.24 | 49 | 163,126 | 30.04 | 54 | 163,760 | 32.98 |
|  | **Total** | 95 | 600,266 | 15.83 | 104 | 614,479 | 16.92 | 107 | 625,404 | 17.11 | 111 | 636,448 | 17.44 |
| **Male** | **26-35** | 6 | 135,281 | 4.44 | 8 | 141,281 | 5.66 | 8 | 146,168 | 5.47 | 2 | 149,788 | 1.34 |
|  | **36-45** | 20 | 151,170 | 13.23 | 17 | 156,808 | 10.84 | 12 | 161,068 | 7.45 | 17 | 164,491 | 10.33 |
|  | **46-55** | 34 | 166,265 | 20.45 | 43 | 170,773 | 25.18 | 39 | 174,180 | 22.39 | 59 | 177,041 | 33.33 |
|  | **56-65** | 69 | 155,750 | 44.3 | 62 | 158,059 | 39.23 | 73 | 158,920 | 45.93 | 77 | 159,711 | 48.21 |
|  | **Total** | 129 | 608,466 | 21.2 | 130 | 626,922 | 20.74 | 132 | 640,337 | 20.61 | 155 | 651,030 | 23.81 |
| **Total** | **26-35** | 13 | 262,889 | 4.95 | 18 | 273,182 | 6.59 | 13 | 281,844 | 4.61 | 7 | 289,412 | 2.42 |
|  | **36-45** | 39 | 297,711 | 13.1 | 39 | 307,279 | 12.69 | 31 | 314,981 | 9.84 | 37 | 321,979 | 11.49 |
|  | **46-55** | 63 | 332,161 | 18.97 | 74 | 340,425 | 21.74 | 73 | 346,869 | 21.05 | 91 | 352,615 | 25.81 |
|  | **56-65** | 109 | 315,970 | 34.5 | 103 | 320,514 | 32.14 | 122 | 322,046 | 37.88 | 131 | 323,471 | 40.5 |
|  | **Total** | 224 | 1,208,732 | 18.53 | 234 | 1,241,401 | 18.85 | 239 | 1,265,740 | 18.88 | 266 | 1,287,478 | 20.66 |

**QUARTERLY INCIDENCE RATES: DVT with anticoagulant 2017 (Cases per 100,000 Person-Years)**

|  |  | **2017 Q1** | | | **2017 Q2** | | | **2017 Q3** | | | **2017 Q4** | | |
| --- | --- | --- | --- | --- | --- | --- | --- | --- | --- | --- | --- | --- | --- |
|  | **Age Group** | **Cases** | **Person-Years** | **Incidence** | **Cases** | **Person-Years** | **Incidence** | **Cases** | **Person-Years** | **Incidence** | **Cases** | **Person-Years** | **Incidence** |
| **Female** | **26-35** | 10 | 135,069 | 7.4 | 7 | 136,727 | 5.12 | 5 | 141,118 | 3.54 | 5 | 145,171 | 3.44 |
|  | **36-45** | 21 | 152,365 | 13.78 | 12 | 153,943 | 7.8 | 20 | 158,025 | 12.66 | 13 | 161,640 | 8.04 |
|  | **46-55** | 23 | 170,401 | 13.5 | 22 | 171,848 | 12.8 | 35 | 176,022 | 19.88 | 27 | 179,627 | 15.03 |
|  | **56-65** | 49 | 171,036 | 28.65 | 35 | 171,066 | 20.46 | 37 | 173,391 | 21.34 | 49 | 174,773 | 28.04 |
|  | **Total** | 103 | 628,872 | 16.38 | 76 | 633,584 | 12 | 97 | 648,556 | 14.96 | 94 | 661,210 | 14.22 |
| **Male** | **26-35** | 9 | 146,711 | 6.13 | 5 | 149,074 | 3.35 | 6 | 153,719 | 3.9 | 2 | 157,833 | 1.27 |
|  | **36-45** | 13 | 160,277 | 8.11 | 18 | 162,703 | 11.06 | 17 | 167,388 | 10.16 | 19 | 171,398 | 11.09 |
|  | **46-55** | 47 | 173,170 | 27.14 | 29 | 175,045 | 16.57 | 34 | 179,515 | 18.94 | 36 | 183,439 | 19.63 |
|  | **56-65** | 55 | 167,493 | 32.84 | 60 | 167,955 | 35.72 | 55 | 170,580 | 32.24 | 57 | 172,342 | 33.07 |
|  | **Total** | 124 | 647,651 | 19.15 | 112 | 654,778 | 17.11 | 112 | 671,202 | 16.69 | 114 | 685,012 | 16.64 |
| **Total** | **26-35** | 19 | 281,781 | 6.74 | 12 | 285,801 | 4.2 | 11 | 294,837 | 3.73 | 7 | 303,004 | 2.31 |
|  | **36-45** | 34 | 312,642 | 10.88 | 30 | 316,646 | 9.47 | 37 | 325,414 | 11.37 | 32 | 333,037 | 9.61 |
|  | **46-55** | 70 | 343,571 | 20.37 | 51 | 346,894 | 14.7 | 69 | 355,536 | 19.41 | 63 | 363,067 | 17.35 |
|  | **56-65** | 104 | 338,529 | 30.72 | 95 | 339,021 | 28.02 | 92 | 343,972 | 26.75 | 106 | 347,115 | 30.54 |
|  | **Total** | 227 | 1,276,523 | 17.78 | 188 | 1,288,362 | 14.59 | 209 | 1,319,759 | 15.84 | 208 | 1,346,222 | 15.45 |

**QUARTERLY INCIDENCE RATES: DVT with anticoagulant 2018 (Cases per 100,000 Person-Years)**

|  |  | **2018 Q1** | | | **2018 Q2** | | | **2018 Q3** | | | **2018 Q4** | | |
| --- | --- | --- | --- | --- | --- | --- | --- | --- | --- | --- | --- | --- | --- |
|  | **Age Group** | **Cases** | **Person-Years** | **Incidence** | **Cases** | **Person-Years** | **Incidence** | **Cases** | **Person-Years** | **Incidence** | **Cases** | **Person-Years** | **Incidence** |
| **Female** | **26-35** | 8 | 142,791 | 5.6 | 5 | 145,165 | 3.44 | 5 | 146,802 | 3.41 | 7 | 148,067 | 4.73 |
|  | **36-45** | 13 | 160,730 | 8.09 | 18 | 163,663 | 11 | 7 | 165,715 | 4.22 | 17 | 166,883 | 10.19 |
|  | **46-55** | 26 | 175,491 | 14.82 | 24 | 178,460 | 13.45 | 33 | 180,708 | 18.26 | 26 | 181,889 | 14.29 |
|  | **56-65** | 38 | 182,781 | 20.79 | 40 | 184,417 | 21.69 | 39 | 185,363 | 21.04 | 26 | 185,060 | 14.05 |
|  | **Total** | 85 | 661,794 | 12.84 | 87 | 671,706 | 12.95 | 84 | 678,588 | 12.38 | 76 | 681,898 | 11.15 |
| **Male** | **26-35** | 2 | 156,818 | 1.28 | 8 | 159,226 | 5.02 | 6 | 161,213 | 3.72 | 7 | 162,557 | 4.31 |
|  | **36-45** | 11 | 171,962 | 6.4 | 11 | 174,725 | 6.3 | 16 | 176,970 | 9.04 | 9 | 178,049 | 5.05 |
|  | **46-55** | 40 | 181,072 | 22.09 | 24 | 183,862 | 13.05 | 30 | 186,258 | 16.11 | 30 | 187,347 | 16.01 |
|  | **56-65** | 57 | 181,405 | 31.42 | 57 | 182,927 | 31.16 | 64 | 183,982 | 34.79 | 62 | 183,680 | 33.75 |
|  | **Total** | 110 | 691,257 | 15.91 | 100 | 700,740 | 14.27 | 116 | 708,423 | 16.37 | 108 | 711,633 | 15.18 |
| **Total** | **26-35** | 10 | 299,609 | 3.34 | 13 | 304,390 | 4.27 | 11 | 308,015 | 3.57 | 14 | 310,624 | 4.51 |
|  | **36-45** | 24 | 332,693 | 7.21 | 29 | 338,388 | 8.57 | 23 | 342,684 | 6.71 | 26 | 344,932 | 7.54 |
|  | **46-55** | 66 | 356,563 | 18.51 | 48 | 362,323 | 13.25 | 63 | 366,965 | 17.17 | 56 | 369,235 | 15.17 |
|  | **56-65** | 95 | 364,186 | 26.09 | 97 | 367,345 | 26.41 | 103 | 369,346 | 27.89 | 88 | 368,740 | 23.87 |
|  | **Total** | 195 | 1,353,051 | 14.41 | 187 | 1,372,446 | 13.63 | 200 | 1,387,011 | 14.42 | 184 | 1,393,531 | 13.2 |

**QUARTERLY INCIDENCE RATES: DVT 2019 with anticoagulant (Cases per 100,000 Person-Years)**

|  |  | **2019 Q1** | | | **2019 Q2** | | | **2019 Q3** | | | **2019 Q4** | | |
| --- | --- | --- | --- | --- | --- | --- | --- | --- | --- | --- | --- | --- | --- |
|  | **Age Group** | **Cases** | **Person-Years** | **Incidence** | **Cases** | **Person-Years** | **Incidence** | **Cases** | **Person-Years** | **Incidence** | **Cases** | **Person-Years** | **Incidence** |
| **Female** | **26-35** | 3 | 143,318 | 2.09 | 1 | 145,559 | 0.69 | 3 | 147,647 | 2.03 | 5 | 149,191 | 3.35 |
|  | **36-45** | 10 | 162,889 | 6.14 | 13 | 165,359 | 7.86 | 4 | 167,453 | 2.39 | 13 | 168,233 | 7.73 |
|  | **46-55** | 19 | 174,683 | 10.88 | 17 | 177,042 | 9.6 | 19 | 179,088 | 10.61 | 15 | 179,894 | 8.34 |
|  | **56-65** | 32 | 188,336 | 16.99 | 26 | 189,396 | 13.73 | 32 | 190,413 | 16.81 | 38 | 189,806 | 20.02 |
|  | **Total** | 64 | 669,226 | 9.56 | 57 | 677,356 | 8.42 | 58 | 684,601 | 8.47 | 71 | 687,124 | 10.33 |
| **Male** | **26-35** | 4 | 157,451 | 2.54 | 4 | 159,569 | 2.51 | 6 | 161,206 | 3.72 | 3 | 162,253 | 1.85 |
|  | **36-45** | 11 | 173,897 | 6.33 | 9 | 176,146 | 5.11 | 6 | 177,668 | 3.38 | 6 | 178,207 | 3.37 |
|  | **46-55** | 24 | 180,199 | 13.32 | 22 | 182,480 | 12.06 | 22 | 184,334 | 11.93 | 22 | 184,919 | 11.9 |
|  | **56-65** | 49 | 186,044 | 26.34 | 51 | 187,101 | 27.26 | 48 | 188,106 | 25.52 | 50 | 187,516 | 26.66 |
|  | **Total** | 88 | 697,591 | 12.61 | 86 | 705,296 | 12.19 | 82 | 711,314 | 11.53 | 81 | 712,895 | 11.36 |
| **Total** | **26-35** | 7 | 300,769 | 2.33 | 5 | 305,128 | 1.64 | 9 | 308,852 | 2.91 | 8 | 311,445 | 2.57 |
|  | **36-45** | 21 | 336,786 | 6.24 | 22 | 341,505 | 6.44 | 10 | 345,121 | 2.9 | 19 | 346,440 | 5.48 |
|  | **46-55** | 43 | 354,882 | 12.12 | 39 | 359,522 | 10.85 | 41 | 363,423 | 11.28 | 37 | 364,812 | 10.14 |
|  | **56-65** | 81 | 374,380 | 21.64 | 77 | 376,497 | 20.45 | 80 | 378,519 | 21.14 | 88 | 377,323 | 23.32 |
|  | **Total** | 152 | 1,366,817 | 11.12 | 143 | 1,382,652 | 10.34 | 140 | 1,395,915 | 10.03 | 152 | 1,400,019 | 10.86 |

**QUARTERLY INCIDENCE RATES: DVT with anticoagulant first quarter 2020 (Cases per 100,000 Person-Years)**

|  |  | **2020 Q1** | | |
| --- | --- | --- | --- | --- |
|  | **Age Group** | **Cases** | **Person-Years** | **Incidence** |
| **Female** | **26-35** | 5 | 139,771 | 3.58 |
|  | **36-45** | 9 | 159,296 | 5.65 |
|  | **46-55** | 21 | 168,201 | 12.49 |
|  | **56-65** | 28 | 188,916 | 14.82 |
|  | **Total** | 63 | 656,184 | 9.6 |
| **Male** | **26-35** | 4 | 152,224 | 2.63 |
|  | **36-45** | 6 | 169,383 | 3.54 |
|  | **46-55** | 24 | 173,832 | 13.81 |
|  | **56-65** | 35 | 186,455 | 18.77 |
|  | **Total** | 69 | 681,895 | 10.12 |
| **Total** | **26-35** | 9 | 291,996 | 3.08 |
|  | **36-45** | 15 | 328,679 | 4.56 |
|  | **46-55** | 45 | 342,033 | 13.16 |
|  | **56-65** | 63 | 375,372 | 16.78 |
|  | **Total** | 132 | 1,338,079 | 9.86 |

**QUARTERLY INCIDENCE RATES: PE with anticoagulant 2015 (**Cases per 100,000 Person-Years)

|  |  | **2015 Q1** | | | **2015 Q2** | | | **2015 Q3** | | | **2015 Q4** | | |
| --- | --- | --- | --- | --- | --- | --- | --- | --- | --- | --- | --- | --- | --- |
|  | **Age Group** | **Cases** | **Person-Years** | **Incidence** | **Cases** | **Person-Years** | **Incidence** | **Cases** | **Person-Years** | **Incidence** | **Cases** | **Person-Years** | **Incidence** |
| **Female** | **26-35** | 70 | 121,625 | 57.55 | 55 | 123,810 | 44.42 | 76 | 125,309 | 60.65 | 54 | 126,514 | 42.68 |
|  | **36-45** | 109 | 140,289 | 77.7 | 95 | 142,992 | 66.44 | 115 | 144,611 | 79.52 | 133 | 145,586 | 91.35 |
|  | **46-55** | 164 | 157,437 | 104.17 | 173 | 160,066 | 108.08 | 181 | 161,590 | 112.01 | 199 | 162,261 | 122.64 |
|  | **56-65** | 230 | 145,538 | 158.03 | 228 | 146,319 | 155.82 | 271 | 145,825 | 185.84 | 288 | 144,535 | 199.26 |
|  | **Total** | 573 | 564,889 | 101.44 | 551 | 573,188 | 96.13 | 643 | 577,335 | 111.37 | 674 | 578,895 | 116.43 |
| **Male** | **26-35** | 34 | 126,557 | 26.87 | 19 | 129,335 | 14.69 | 31 | 131,713 | 23.54 | 27 | 132,662 | 20.35 |
|  | **36-45** | 85 | 144,268 | 58.92 | 85 | 147,430 | 57.65 | 106 | 149,704 | 70.81 | 103 | 150,443 | 68.46 |
|  | **46-55** | 211 | 157,837 | 133.68 | 174 | 160,937 | 108.12 | 199 | 162,868 | 122.18 | 239 | 163,209 | 146.44 |
|  | **56-65** | 364 | 142,437 | 255.55 | 324 | 143,492 | 225.8 | 344 | 143,621 | 239.52 | 354 | 142,439 | 248.53 |
|  | **Total** | 694 | 571,100 | 121.52 | 602 | 581,193 | 103.58 | 680 | 587,906 | 115.66 | 723 | 588,752 | 122.8 |
| **Total** | **26-35** | 104 | 248,182 | 41.9 | 74 | 253,145 | 29.23 | 107 | 257,022 | 41.63 | 81 | 259,176 | 31.25 |
|  | **36-45** | 194 | 284,557 | 68.18 | 180 | 290,422 | 61.98 | 221 | 294,315 | 75.09 | 236 | 296,029 | 79.72 |
|  | **46-55** | 375 | 315,275 | 118.94 | 347 | 321,003 | 108.1 | 380 | 324,458 | 117.12 | 438 | 325,470 | 134.57 |
|  | **56-65** | 594 | 287,976 | 206.27 | 552 | 289,812 | 190.47 | 615 | 289,446 | 212.48 | 642 | 286,973 | 223.71 |
|  | **Total** | 1,267 | 1,135,989 | 111.53 | 1,153 | 1,154,381 | 99.88 | 1,323 | 1,165,241 | 113.54 | 1,397 | 1,167,648 | 119.64 |
| CASE is PE with or without DVT AND anticoagulant Rx within 30d OR disenrollment in 30d | | | | | | | | |  |  |  |  |  |

**QUARTERLY INCIDENCE RATES: PE with anticoagulant 2016 (**Cases per 100,000 Person-Years)

|  |  | **2016 Q1** | | | **2016 Q2** | | | **2016 Q3** | | | **2016 Q4** | | |
| --- | --- | --- | --- | --- | --- | --- | --- | --- | --- | --- | --- | --- | --- |
|  | **Age Group** | **Cases** | **Person-Years** | **Incidence** | **Cases** | **Person-Years** | **Incidence** | **Cases** | **Person-Years** | **Incidence** | **Cases** | **Person-Years** | **Incidence** |
| **Female** | **26-35** | 47 | 127,542 | 36.85 | 53 | 131,831 | 40.2 | 67 | 135,602 | 49.41 | 74 | 139,540 | 53.03 |
|  | **36-45** | 111 | 146,386 | 75.83 | 113 | 150,309 | 75.18 | 117 | 153,749 | 76.1 | 136 | 157,317 | 86.45 |
|  | **46-55** | 178 | 165,639 | 107.46 | 174 | 169,388 | 102.72 | 197 | 172,412 | 114.26 | 206 | 175,293 | 117.52 |
|  | **56-65** | 300 | 159,827 | 187.7 | 275 | 162,043 | 169.71 | 282 | 162,709 | 173.32 | 335 | 163,336 | 205.1 |
|  | **Total** | 636 | 599,393 | 106.11 | 615 | 613,571 | 100.23 | 663 | 624,471 | 106.17 | 751 | 635,486 | 118.18 |
| **Male** | **26-35** | 23 | 135,246 | 17.01 | 36 | 141,244 | 25.49 | 34 | 146,128 | 23.27 | 41 | 149,743 | 27.38 |
|  | **36-45** | 87 | 151,057 | 57.59 | 88 | 156,693 | 56.16 | 91 | 160,943 | 56.54 | 93 | 164,363 | 56.58 |
|  | **46-55** | 202 | 166,011 | 121.68 | 216 | 170,503 | 126.68 | 239 | 173,890 | 137.44 | 269 | 176,738 | 152.2 |
|  | **56-65** | 403 | 155,270 | 259.55 | 400 | 157,546 | 253.89 | 404 | 158,392 | 255.06 | 432 | 159,171 | 271.41 |
|  | **Total** | 715 | 607,584 | 117.68 | 740 | 625,986 | 118.21 | 768 | 639,353 | 120.12 | 835 | 650,015 | 128.46 |
| **Total** | **26-35** | 70 | 262,788 | 26.64 | 89 | 273,075 | 32.59 | 101 | 281,730 | 35.85 | 115 | 289,282 | 39.75 |
|  | **36-45** | 198 | 297,444 | 66.57 | 201 | 307,002 | 65.47 | 208 | 314,691 | 66.1 | 229 | 321,680 | 71.19 |
|  | **46-55** | 380 | 331,649 | 114.58 | 390 | 339,891 | 114.74 | 436 | 346,303 | 125.9 | 475 | 352,031 | 134.93 |
|  | **56-65** | 703 | 315,096 | 223.11 | 675 | 319,588 | 211.21 | 686 | 321,101 | 213.64 | 767 | 322,507 | 237.82 |
|  | **Total** | 1,351 | 1,206,977 | 111.93 | 1,355 | 1,239,556 | 109.31 | 1,431 | 1,263,825 | 113.23 | 1,586 | 1,285,501 | 123.38 |
| CASE is PE with or without DVT AND anticoagulant Rx within 30d OR disenrollment in 30d | | | | | | | | |  |  |  |  |  |

**QUARTERLY INCIDENCE RATES: PE with anticoagulant 2017 (**Cases per 100,000 Person-Years)

|  |  | **2017 Q1** | | | **2017 Q2** | | | **2017 Q3** | | | **2017 Q4** | | |
| --- | --- | --- | --- | --- | --- | --- | --- | --- | --- | --- | --- | --- | --- |
|  | **Age Group** | **Cases** | **Person-Years** | **Incidence** | **Cases** | **Person-Years** | **Incidence** | **Cases** | **Person-Years** | **Incidence** | **Cases** | **Person-Years** | **Incidence** |
| **Female** | **26-35** | 58 | 134,991 | 42.97 | 54 | 136,645 | 39.52 | 67 | 141,033 | 47.51 | 52 | 145,076 | 35.84 |
|  | **36-45** | 136 | 152,200 | 89.36 | 121 | 153,769 | 78.69 | 136 | 157,845 | 86.16 | 133 | 161,456 | 82.38 |
|  | **46-55** | 224 | 170,092 | 131.69 | 190 | 171,534 | 110.76 | 179 | 175,700 | 101.88 | 217 | 179,302 | 121.02 |
|  | **56-65** | 365 | 170,576 | 213.98 | 340 | 170,594 | 199.3 | 348 | 172,903 | 201.27 | 415 | 174,259 | 238.15 |
|  | **Total** | 783 | 627,859 | 124.71 | 705 | 632,542 | 111.46 | 730 | 647,481 | 112.74 | 817 | 660,093 | 123.77 |
| **Male** | **26-35** | 24 | 146,672 | 16.36 | 35 | 149,033 | 23.48 | 35 | 153,676 | 22.78 | 46 | 157,786 | 29.15 |
|  | **36-45** | 102 | 160,151 | 63.69 | 104 | 162,573 | 63.97 | 90 | 167,252 | 53.81 | 98 | 171,258 | 57.22 |
|  | **46-55** | 244 | 172,880 | 141.14 | 221 | 174,744 | 126.47 | 237 | 179,199 | 132.26 | 237 | 183,107 | 129.43 |
|  | **56-65** | 415 | 166,929 | 248.61 | 430 | 167,379 | 256.9 | 449 | 169,981 | 264.15 | 489 | 171,727 | 284.75 |
|  | **Total** | 785 | 646,633 | 121.4 | 790 | 653,729 | 120.85 | 811 | 670,108 | 121.03 | 870 | 683,879 | 127.22 |
| **Total** | **26-35** | 82 | 281,663 | 29.11 | 89 | 285,677 | 31.15 | 102 | 294,709 | 34.61 | 98 | 302,862 | 32.36 |
|  | **36-45** | 238 | 312,352 | 76.2 | 225 | 316,342 | 71.13 | 226 | 325,097 | 69.52 | 231 | 332,714 | 69.43 |
|  | **46-55** | 468 | 342,971 | 136.45 | 411 | 346,279 | 118.69 | 416 | 354,899 | 117.22 | 454 | 362,409 | 125.27 |
|  | **56-65** | 780 | 337,505 | 231.11 | 770 | 337,973 | 227.83 | 797 | 342,883 | 232.44 | 904 | 345,987 | 261.28 |
|  | **Total** | 1,568 | 1,274,492 | 123.03 | 1,495 | 1,286,271 | 116.23 | 1,541 | 1,317,588 | 116.96 | 1,687 | 1,343,972 | 125.52 |
| CASE is PE with or without DVT AND anticoagulant Rx within 30d OR disenrollment in 30d | | | | | | | | |  |  |  |  |  |

**QUARTERLY INCIDENCE RATES: PE with anticoagulant 2018 (**Cases per 100,000 Person-Years)

|  |  | **2018 Q1** | | | **2018 Q2** | | | **2018 Q3** | | | **2018 Q4** | | |
| --- | --- | --- | --- | --- | --- | --- | --- | --- | --- | --- | --- | --- | --- |
|  | **Age Group** | **Cases** | **Person-Years** | **Incidence** | **Cases** | **Person-Years** | **Incidence** | **Cases** | **Person-Years** | **Incidence** | **Cases** | **Person-Years** | **Incidence** |
| **Female** | **26-35** | 53 | 142,705 | 37.14 | 57 | 145,079 | 39.29 | 71 | 146,717 | 48.39 | 75 | 147,974 | 50.68 |
|  | **36-45** | 128 | 160,561 | 79.72 | 130 | 163,486 | 79.52 | 136 | 165,528 | 82.16 | 149 | 166,686 | 89.39 |
|  | **46-55** | 229 | 175,192 | 130.71 | 225 | 178,144 | 126.3 | 261 | 180,382 | 144.69 | 244 | 181,543 | 134.4 |
|  | **56-65** | 385 | 182,248 | 211.25 | 407 | 183,858 | 221.37 | 401 | 184,789 | 217 | 449 | 184,454 | 243.42 |
|  | **Total** | 795 | 660,706 | 120.33 | 819 | 670,568 | 122.14 | 869 | 677,415 | 128.28 | 917 | 680,656 | 134.72 |
| **Male** | **26-35** | 47 | 156,776 | 29.98 | 44 | 159,179 | 27.64 | 47 | 161,159 | 29.16 | 44 | 162,495 | 27.08 |
|  | **36-45** | 101 | 171,832 | 58.78 | 96 | 174,586 | 54.99 | 106 | 176,825 | 59.95 | 100 | 177,895 | 56.21 |
|  | **46-55** | 254 | 180,765 | 140.51 | 242 | 183,543 | 131.85 | 250 | 185,928 | 134.46 | 293 | 186,997 | 156.69 |
|  | **56-65** | 480 | 180,766 | 265.54 | 468 | 182,259 | 256.78 | 497 | 183,296 | 271.15 | 552 | 182,976 | 301.68 |
|  | **Total** | 882 | 690,138 | 127.8 | 850 | 699,567 | 121.5 | 900 | 707,208 | 127.26 | 989 | 710,363 | 139.22 |
| **Total** | **26-35** | 100 | 299,481 | 33.39 | 101 | 304,258 | 33.2 | 118 | 307,876 | 38.33 | 119 | 310,469 | 38.33 |
|  | **36-45** | 229 | 332,393 | 68.89 | 226 | 338,072 | 66.85 | 242 | 342,353 | 70.69 | 249 | 344,580 | 72.26 |
|  | **46-55** | 483 | 355,957 | 135.69 | 467 | 361,687 | 129.12 | 511 | 366,309 | 139.5 | 537 | 368,539 | 145.71 |
|  | **56-65** | 865 | 363,014 | 238.28 | 875 | 366,117 | 238.99 | 898 | 368,085 | 243.97 | 1,001 | 367,430 | 272.43 |
|  | **Total** | 1,677 | 1,350,845 | 124.14 | 1,669 | 1,370,134 | 121.81 | 1,769 | 1,384,623 | 127.76 | 1,906 | 1,391,019 | 137.02 |
| CASE is PE with or without DVT AND anticoagulant Rx within 30d OR disenrollment in 30d | | | | | | | | |  |  |  |  |  |

**QUARTERLY INCIDENCE RATES: PE with anticoagulant 2019 (**Cases per 100,000 Person-Years)

|  |  | **2019 Q1** | | | **2019 Q2** | | | **2019 Q3** | | | **2019 Q4** | | |
| --- | --- | --- | --- | --- | --- | --- | --- | --- | --- | --- | --- | --- | --- |
|  | **Age Group** | **Cases** | **Person-Years** | **Incidence** | **Cases** | **Person-Years** | **Incidence** | **Cases** | **Person-Years** | **Incidence** | **Cases** | **Person-Years** | **Incidence** |
| **Female** | **26-35** | 69 | 143,226 | 48.18 | 75 | 145,463 | 51.56 | 69 | 147,540 | 46.77 | 79 | 149,082 | 52.99 |
|  | **36-45** | 135 | 162,702 | 82.97 | 144 | 165,163 | 87.19 | 166 | 167,248 | 99.25 | 152 | 168,014 | 90.47 |
|  | **46-55** | 237 | 174,339 | 135.94 | 243 | 176,678 | 137.54 | 239 | 178,711 | 133.74 | 251 | 179,504 | 139.83 |
|  | **56-65** | 435 | 187,747 | 231.7 | 414 | 188,788 | 219.29 | 434 | 189,788 | 228.68 | 433 | 189,168 | 228.9 |
|  | **Total** | 876 | 668,014 | 131.13 | 876 | 676,092 | 129.57 | 908 | 683,288 | 132.89 | 915 | 685,769 | 133.43 |
| **Male** | **26-35** | 44 | 157,398 | 27.95 | 43 | 159,518 | 26.96 | 54 | 161,151 | 33.51 | 49 | 162,194 | 30.21 |
|  | **36-45** | 90 | 173,761 | 51.8 | 98 | 176,004 | 55.68 | 126 | 177,516 | 70.98 | 132 | 178,045 | 74.14 |
|  | **46-55** | 242 | 179,878 | 134.54 | 223 | 182,135 | 122.44 | 226 | 183,981 | 122.84 | 283 | 184,557 | 153.34 |
|  | **56-65** | 524 | 185,345 | 282.72 | 477 | 186,406 | 255.89 | 543 | 187,393 | 289.77 | 557 | 186,774 | 298.22 |
|  | **Total** | 900 | 696,383 | 129.24 | 841 | 704,063 | 119.45 | 949 | 710,042 | 133.65 | 1,021 | 711,570 | 143.49 |
| **Total** | **26-35** | 113 | 300,625 | 37.59 | 118 | 304,981 | 38.69 | 123 | 308,692 | 39.85 | 128 | 311,277 | 41.12 |
|  | **36-45** | 225 | 336,463 | 66.87 | 242 | 341,167 | 70.93 | 292 | 344,764 | 84.7 | 284 | 346,059 | 82.07 |
|  | **46-55** | 479 | 354,217 | 135.23 | 466 | 358,813 | 129.87 | 465 | 362,693 | 128.21 | 534 | 364,061 | 146.68 |
|  | **56-65** | 959 | 373,092 | 257.04 | 891 | 375,194 | 237.48 | 977 | 377,181 | 259.03 | 990 | 375,942 | 263.34 |
|  | **Total** | 1,776 | 1,364,397 | 130.17 | 1,717 | 1,380,155 | 124.41 | 1,857 | 1,393,330 | 133.28 | 1,936 | 1,397,339 | 138.55 |

CASE is PE with or without DVT AND anticoagulant Rx within 30d OR disenrollment in 30d

**QUARTERLY INCIDENCE RATES: PE with anticoagulant first quarter 2020 (Cases per 100,000 Person-Years)**

|  |  | **2020 Q1** | | |
| --- | --- | --- | --- | --- |
|  | **Age Group** | **Cases** | **Person-Years** | **Incidence** |
| **Female** | **26-35** | 72 | 139,682 | 51.55 |
|  | **36-45** | 147 | 159,096 | 92.4 |
|  | **46-55** | 239 | 167,842 | 142.4 |
|  | **56-65** | 406 | 188,284 | 215.63 |
|  | **Total** | 864 | 654,904 | 131.93 |
| **Male** | **26-35** | 48 | 152,168 | 31.54 |
|  | **36-45** | 111 | 169,235 | 65.59 |
|  | **46-55** | 272 | 173,493 | 156.78 |
|  | **56-65** | 561 | 185,727 | 302.06 |
|  | **Total** | 992 | 680,623 | 145.75 |
| **Total** | **26-35** | 120 | 291,850 | 41.12 |
|  | **36-45** | 258 | 328,332 | 78.58 |
|  | **46-55** | 511 | 341,335 | 149.71 |
|  | **56-65** | 967 | 374,011 | 258.55 |
|  | **Total** | 1,856 | 1,335,527 | 138.97 |

CASE is PE with or without DVT AND anticoagulant Rx within 30d OR disenrollment in 30d

**QUARTERLY INCIDENCE RATES: VTE with anticoagulant 2015 (**Cases per 100,000 Person-Years)

|  |  | **2015 Q1** | | | **2015 Q2** | | | **2015 Q3** | | | **2015 Q4** | | |
| --- | --- | --- | --- | --- | --- | --- | --- | --- | --- | --- | --- | --- | --- |
|  | **Age Group** | **Cases** | **Person-Years** | **Incidence** | **Cases** | **Person-Years** | **Incidence** | **Cases** | **Person-Years** | **Incidence** | **Cases** | **Person-Years** | **Incidence** |
| **Female** | **26-35** | 84 | 121,606 | 69.08 | 69 | 123,792 | 55.74 | 89 | 125,295 | 71.03 | 63 | 126,499 | 49.8 |
|  | **36-45** | 133 | 140,251 | 94.83 | 107 | 142,955 | 74.85 | 142 | 144,575 | 98.22 | 148 | 145,553 | 101.68 |
|  | **46-55** | 208 | 157,358 | 132.18 | 212 | 159,989 | 132.51 | 224 | 161,517 | 138.69 | 228 | 162,190 | 140.58 |
|  | **56-65** | 282 | 145,435 | 193.9 | 284 | 146,217 | 194.23 | 331 | 145,723 | 227.14 | 326 | 144,439 | 225.7 |
|  | **Total** | 707 | 564,650 | 125.21 | 672 | 572,952 | 117.29 | 786 | 577,109 | 136.2 | 765 | 578,682 | 132.2 |
| **Male** | **26-35** | 45 | 126,543 | 35.56 | 25 | 129,320 | 19.33 | 43 | 131,698 | 32.65 | 32 | 132,647 | 24.12 |
|  | **36-45** | 109 | 144,227 | 75.58 | 100 | 147,390 | 67.85 | 121 | 149,668 | 80.85 | 119 | 150,409 | 79.12 |
|  | **46-55** | 277 | 157,739 | 175.61 | 232 | 160,836 | 144.25 | 246 | 162,770 | 151.13 | 273 | 163,114 | 167.37 |
|  | **56-65** | 419 | 142,278 | 294.49 | 413 | 143,338 | 288.13 | 416 | 143,475 | 289.95 | 430 | 142,300 | 302.18 |
|  | **Total** | 850 | 570,786 | 148.92 | 770 | 580,884 | 132.56 | 826 | 587,610 | 140.57 | 854 | 588,469 | 145.12 |
| **Total** | **26-35** | 129 | 248,148 | 51.99 | 94 | 253,111 | 37.14 | 132 | 256,993 | 51.36 | 95 | 259,146 | 36.66 |
|  | **36-45** | 242 | 284,478 | 85.07 | 207 | 290,345 | 71.29 | 263 | 294,243 | 89.38 | 267 | 295,962 | 90.21 |
|  | **46-55** | 485 | 315,097 | 153.92 | 444 | 320,825 | 138.39 | 470 | 324,287 | 144.93 | 501 | 325,304 | 154.01 |
|  | **56-65** | 701 | 287,713 | 243.65 | 697 | 289,554 | 240.71 | 747 | 289,197 | 258.3 | 756 | 286,739 | 263.65 |
|  | **Total** | 1,557 | 1,135,436 | 137.13 | 1,442 | 1,153,836 | 124.97 | 1,612 | 1,164,720 | 138.4 | 1,619 | 1,167,151 | 138.71 |
| CASE DVT or PE AND anticoagulant Rx within 30d OR disenrollment in 30d | | | | | | | |  |  |  |  |  |  |

**QUARTERLY INCIDENCE RATES: VTE with anticoagulant 2016 (**Cases per 100,000 Person-Years)

|  |  | **2016 Q1** | | | **2016 Q2** | | | **2016 Q3** | | | **2016 Q4** | | |
| --- | --- | --- | --- | --- | --- | --- | --- | --- | --- | --- | --- | --- | --- |
|  | **Age Group** | **Cases** | **Person-Years** | **Incidence** | **Cases** | **Person-Years** | **Incidence** | **Cases** | **Person-Years** | **Incidence** | **Cases** | **Person-Years** | **Incidence** |
| **Female** | **26-35** | 54 | 127,528 | 42.34 | 62 | 131,818 | 47.03 | 72 | 135,589 | 53.1 | 79 | 139,531 | 56.62 |
|  | **36-45** | 130 | 146,357 | 88.82 | 132 | 150,282 | 87.84 | 137 | 153,721 | 89.12 | 155 | 157,293 | 98.54 |
|  | **46-55** | 204 | 165,579 | 123.2 | 203 | 169,328 | 119.89 | 222 | 172,355 | 128.8 | 240 | 175,244 | 136.95 |
|  | **56-65** | 332 | 159,743 | 207.83 | 311 | 161,962 | 192.02 | 325 | 162,632 | 199.84 | 377 | 163,271 | 230.9 |
|  | **Total** | 720 | 599,207 | 120.16 | 708 | 613,390 | 115.42 | 756 | 624,298 | 121.1 | 851 | 635,338 | 133.94 |
| **Male** | **26-35** | 29 | 135,234 | 21.44 | 44 | 141,234 | 31.15 | 43 | 146,118 | 29.43 | 42 | 149,735 | 28.05 |
|  | **36-45** | 104 | 151,026 | 68.86 | 104 | 156,666 | 66.38 | 101 | 160,919 | 62.76 | 110 | 164,339 | 66.93 |
|  | **46-55** | 234 | 165,926 | 141.03 | 253 | 170,424 | 148.45 | 276 | 173,817 | 158.79 | 322 | 176,669 | 182.26 |
|  | **56-65** | 465 | 155,129 | 299.75 | 452 | 157,413 | 287.14 | 469 | 158,269 | 296.33 | 507 | 159,059 | 318.75 |
|  | **Total** | 832 | 607,316 | 137 | 853 | 625,736 | 136.32 | 889 | 639,124 | 139.1 | 981 | 649,803 | 150.97 |
| **Total** | **26-35** | 83 | 262,762 | 31.59 | 106 | 273,052 | 38.82 | 115 | 281,708 | 40.82 | 121 | 289,266 | 41.83 |
|  | **36-45** | 234 | 297,383 | 78.69 | 236 | 306,948 | 76.89 | 238 | 314,640 | 75.64 | 265 | 321,633 | 82.39 |
|  | **46-55** | 438 | 331,505 | 132.12 | 456 | 339,751 | 134.22 | 498 | 346,173 | 143.86 | 562 | 351,913 | 159.7 |
|  | **56-65** | 797 | 314,872 | 253.12 | 763 | 319,375 | 238.9 | 794 | 320,901 | 247.43 | 884 | 322,330 | 274.25 |
|  | **Total** | 1,552 | 1,206,523 | 128.63 | 1,561 | 1,239,126 | 125.98 | 1,645 | 1,263,422 | 130.2 | 1,832 | 1,285,141 | 142.55 |
| CASE DVT or PE AND anticoagulant Rx within 30d OR disenrollment in 30d | | | | | | | |  |  |  |  |  |  |

**QUARTERLY INCIDENCE RATES: VTE with anticoagulant 2017 (**Cases per 100,000 Person-Years)

|  |  | **2017 Q1** | | | **2017 Q2** | | | **2017 Q3** | | | **2017 Q4** | | |
| --- | --- | --- | --- | --- | --- | --- | --- | --- | --- | --- | --- | --- | --- |
|  | **Age Group** | **Cases** | **Person-Years** | **Incidence** | **Cases** | **Person-Years** | **Incidence** | **Cases** | **Person-Years** | **Incidence** | **Cases** | **Person-Years** | **Incidence** |
| **Female** | **26-35** | 68 | 134,982 | 50.38 | 61 | 136,637 | 44.64 | 72 | 141,026 | 51.05 | 58 | 145,070 | 39.98 |
|  | **36-45** | 151 | 152,177 | 99.23 | 132 | 153,748 | 85.85 | 156 | 157,823 | 98.84 | 145 | 161,433 | 89.82 |
|  | **46-55** | 243 | 170,049 | 142.9 | 209 | 171,494 | 121.87 | 213 | 175,657 | 121.26 | 243 | 179,259 | 135.56 |
|  | **56-65** | 401 | 170,508 | 235.18 | 367 | 170,528 | 215.21 | 378 | 172,838 | 218.7 | 457 | 174,194 | 262.35 |
|  | **Total** | 863 | 627,716 | 137.48 | 769 | 632,406 | 121.6 | 819 | 647,345 | 126.52 | 903 | 659,955 | 136.83 |
| **Male** | **26-35** | 33 | 146,665 | 22.5 | 39 | 149,025 | 26.17 | 41 | 153,669 | 26.68 | 49 | 157,780 | 31.06 |
|  | **36-45** | 115 | 160,133 | 71.82 | 121 | 162,553 | 74.44 | 106 | 167,231 | 63.39 | 118 | 171,236 | 68.91 |
|  | **46-55** | 288 | 172,816 | 166.65 | 247 | 174,681 | 141.4 | 266 | 179,139 | 148.49 | 270 | 183,050 | 147.5 |
|  | **56-65** | 464 | 166,813 | 278.16 | 479 | 167,271 | 286.36 | 498 | 169,877 | 293.15 | 538 | 171,626 | 313.47 |
|  | **Total** | 900 | 646,428 | 139.23 | 886 | 653,531 | 135.57 | 911 | 669,916 | 135.99 | 975 | 683,691 | 142.61 |
| **Total** | **26-35** | 101 | 281,648 | 35.86 | 100 | 285,662 | 35.01 | 113 | 294,695 | 38.34 | 107 | 302,849 | 35.33 |
|  | **36-45** | 266 | 312,310 | 85.17 | 253 | 316,301 | 79.99 | 262 | 325,055 | 80.6 | 263 | 332,669 | 79.06 |
|  | **46-55** | 531 | 342,865 | 154.87 | 456 | 346,175 | 131.73 | 479 | 354,796 | 135.01 | 513 | 362,308 | 141.59 |
|  | **56-65** | 865 | 337,321 | 256.43 | 846 | 337,799 | 250.44 | 876 | 342,715 | 255.61 | 995 | 345,819 | 287.72 |
|  | **Total** | 1,763 | 1,274,144 | 138.37 | 1,655 | 1,285,937 | 128.7 | 1,730 | 1,317,261 | 131.33 | 1,878 | 1,343,646 | 139.77 |

CASE DVT or PE AND anticoagulant Rx within 30d OR disenrollment in 30d

**QUARTERLY INCIDENCE RATES: VTE with anticoagulant 2018 (**Cases per 100,000 Person-Years)

|  |  | **2018 Q1** | | | **2018 Q2** | | | **2018 Q3** | | | **2018 Q4** | | |
| --- | --- | --- | --- | --- | --- | --- | --- | --- | --- | --- | --- | --- | --- |
|  | **Age Group** | **Cases** | **Person-Years** | **Incidence** | **Cases** | **Person-Years** | **Incidence** | **Cases** | **Person-Years** | **Incidence** | **Cases** | **Person-Years** | **Incidence** |
| **Female** | **26-35** | 60 | 142,698 | 42.05 | 62 | 145,072 | 42.74 | 75 | 146,711 | 51.12 | 82 | 147,967 | 55.42 |
|  | **36-45** | 140 | 160,540 | 87.21 | 147 | 163,465 | 89.93 | 143 | 165,508 | 86.4 | 163 | 166,667 | 97.8 |
|  | **46-55** | 252 | 175,154 | 143.87 | 247 | 178,107 | 138.68 | 288 | 180,343 | 159.7 | 269 | 181,505 | 148.21 |
|  | **56-65** | 418 | 182,188 | 229.43 | 444 | 183,798 | 241.57 | 437 | 184,729 | 236.56 | 469 | 184,398 | 254.34 |
|  | **Total** | 870 | 660,580 | 131.7 | 900 | 670,441 | 134.24 | 943 | 677,290 | 139.23 | 983 | 680,538 | 144.44 |
| **Male** | **26-35** | 48 | 156,769 | 30.62 | 52 | 159,173 | 32.67 | 51 | 161,153 | 31.65 | 50 | 162,489 | 30.77 |
|  | **36-45** | 112 | 171,811 | 65.19 | 105 | 174,566 | 60.15 | 120 | 176,806 | 67.87 | 108 | 177,878 | 60.72 |
|  | **46-55** | 288 | 180,712 | 159.37 | 265 | 183,495 | 144.42 | 278 | 185,880 | 149.56 | 321 | 186,950 | 171.7 |
|  | **56-65** | 532 | 180,673 | 294.46 | 522 | 182,165 | 286.55 | 560 | 183,203 | 305.67 | 611 | 182,883 | 334.09 |
|  | **Total** | 980 | 689,965 | 142.04 | 944 | 699,400 | 134.97 | 1,009 | 707,042 | 142.71 | 1,090 | 710,199 | 153.48 |
| **Total** | **26-35** | 108 | 299,467 | 36.06 | 114 | 304,245 | 37.47 | 126 | 307,864 | 40.93 | 132 | 310,456 | 42.52 |
|  | **36-45** | 252 | 332,352 | 75.82 | 252 | 338,031 | 74.55 | 263 | 342,315 | 76.83 | 271 | 344,545 | 78.65 |
|  | **46-55** | 540 | 355,867 | 151.74 | 512 | 361,602 | 141.59 | 566 | 366,223 | 154.55 | 590 | 368,455 | 160.13 |
|  | **56-65** | 950 | 362,860 | 261.81 | 966 | 365,963 | 263.96 | 997 | 367,931 | 270.97 | 1,080 | 367,281 | 294.05 |
|  | **Total** | 1,850 | 1,350,546 | 136.98 | 1,844 | 1,369,841 | 134.61 | 1,952 | 1,384,332 | 141.01 | 2,073 | 1,390,737 | 149.06 |

CASE DVT or PE AND anticoagulant Rx within 30d OR disenrollment in 30d

**QUARTERLY INCIDENCE RATES: VTE with anticoagulant 2019 (**Cases per 100,000 Person-Years)

|  |  | **2019 Q1** | | | **2019 Q2** | | | **2019 Q3** | | | **2019 Q4** | | |
| --- | --- | --- | --- | --- | --- | --- | --- | --- | --- | --- | --- | --- | --- |
|  | **Age Group** | **Cases** | **Person-Years** | **Incidence** | **Cases** | **Person-Years** | **Incidence** | **Cases** | **Person-Years** | **Incidence** | **Cases** | **Person-Years** | **Incidence** |
| **Female** | **26-35** | 72 | 143,221 | 50.27 | 76 | 145,459 | 52.25 | 72 | 147,537 | 48.8 | 84 | 149,078 | 56.35 |
|  | **36-45** | 144 | 162,688 | 88.51 | 158 | 165,150 | 95.67 | 169 | 167,236 | 101.05 | 165 | 168,004 | 98.21 |
|  | **46-55** | 254 | 174,309 | 145.72 | 260 | 176,647 | 147.19 | 257 | 178,683 | 143.83 | 265 | 179,477 | 147.65 |
|  | **56-65** | 466 | 187,694 | 248.28 | 437 | 188,737 | 231.54 | 466 | 189,739 | 245.6 | 471 | 189,120 | 249.05 |
|  | **Total** | 936 | 667,912 | 140.14 | 931 | 675,993 | 137.72 | 964 | 683,195 | 141.1 | 985 | 685,679 | 143.65 |
| **Male** | **26-35** | 46 | 157,392 | 29.23 | 47 | 159,510 | 29.47 | 59 | 161,143 | 36.61 | 52 | 162,187 | 32.06 |
|  | **36-45** | 101 | 173,748 | 58.13 | 107 | 175,992 | 60.8 | 130 | 177,504 | 73.24 | 137 | 178,034 | 76.95 |
|  | **46-55** | 264 | 179,841 | 146.8 | 243 | 182,103 | 133.44 | 244 | 183,950 | 132.64 | 302 | 184,526 | 163.66 |
|  | **56-65** | 570 | 185,263 | 307.67 | 526 | 186,322 | 282.31 | 590 | 187,311 | 314.98 | 606 | 186,698 | 324.59 |
|  | **Total** | 981 | 696,244 | 140.9 | 923 | 703,927 | 131.12 | 1,023 | 709,907 | 144.1 | 1,097 | 711,446 | 154.19 |
| **Total** | **26-35** | 118 | 300,613 | 39.25 | 123 | 304,969 | 40.33 | 131 | 308,679 | 42.44 | 136 | 311,266 | 43.69 |
|  | **36-45** | 245 | 336,436 | 72.82 | 265 | 341,141 | 77.68 | 299 | 344,740 | 86.73 | 302 | 346,038 | 87.27 |
|  | **46-55** | 518 | 354,149 | 146.27 | 503 | 358,750 | 140.21 | 501 | 362,633 | 138.16 | 567 | 364,003 | 155.77 |
|  | **56-65** | 1,036 | 372,957 | 277.78 | 963 | 375,059 | 256.76 | 1,056 | 377,050 | 280.07 | 1,077 | 375,818 | 286.57 |
|  | **Total** | 1,917 | 1,364,156 | 140.53 | 1,854 | 1,379,919 | 134.36 | 1,987 | 1,393,102 | 142.63 | 2,082 | 1,397,125 | 149.02 |
| CASE DVT or PE AND anticoagulant Rx within 30d OR disenrollment in 30d | | | | | | | |  |  |  |  |  |  |

**QUARTERLY INCIDENCE RATES: VTE with anticoagulant first quarter 2020 (**Cases per 100,000 Person-Years)

|  |  | **2020 Q1** | | |
| --- | --- | --- | --- | --- |
|  | **Age Group** | **Cases** | **Person-Years** | **Incidence** |
| **Female** | **26-35** | 77 | 139,678 | 55.13 |
|  | **36-45** | 156 | 159,087 | 98.06 |
|  | **46-55** | 260 | 167,819 | 154.93 |
|  | **56-65** | 434 | 188,239 | 230.56 |
|  | **Total** | 927 | 654,823 | 141.56 |
| **Male** | **26-35** | 52 | 152,164 | 34.17 |
|  | **36-45** | 119 | 169,224 | 70.32 |
|  | **46-55** | 296 | 173,467 | 170.64 |
|  | **56-65** | 596 | 185,658 | 321.02 |
|  | **Total** | 1,063 | 680,513 | 156.21 |
| **Total** | **26-35** | 129 | 291,842 | 44.2 |
|  | **36-45** | 275 | 328,311 | 83.76 |
|  | **46-55** | 556 | 341,286 | 162.91 |
|  | **56-65** | 1,030 | 373,897 | 275.48 |
|  | **Total** | 1,990 | 1,335,336 | 149.03 |

CASE DVT or PE AND anticoagulant Rx within 30d OR disenrollment in 30d

**QUARTERLY INCIDENCE RATES: CVT 2015 (Cases per 100,000 Person-Years)**

|  |  | **2015 Q1** | | | **2015 Q2** | | | **2015 Q3** | | | **2015 Q4** | | |
| --- | --- | --- | --- | --- | --- | --- | --- | --- | --- | --- | --- | --- | --- |
|  | **Age Group** | **Cases** | **Person-Years** | **Incidence** | **Cases** | **Person-Years** | **Incidence** | **Cases** | **Person-Years** | **Incidence** | **Cases** | **Person-Years** | **Incidence** |
| **Female** | **26-35** | 3 | 121,698 | 2.47 | 7 | 123,888 | 5.65 | 7 | 125,390 | 5.58 | 5 | 126,598 | 3.95 |
|  | **36-45** | 4 | 140,443 | 2.85 | 11 | 143,149 | 7.68 | 9 | 144,770 | 6.22 | 13 | 145,755 | 8.92 |
|  | **46-55** | 16 | 157,708 | 10.15 | 12 | 160,345 | 7.48 | 12 | 161,876 | 7.41 | 11 | 162,551 | 6.77 |
|  | **56-65** | 3 | 145,976 | 2.06 | 10 | 146,759 | 6.81 | 5 | 146,261 | 3.42 | 21 | 144,971 | 14.49 |
|  | **Total** | 26 | 565,825 | 4.6 | 40 | 574,141 | 6.97 | 33 | 578,298 | 5.71 | 50 | 579,875 | 8.62 |
| **Male** | **26-35** | 1 | 126,599 | 0.79 | 0 | 129,378 | 0 | 3 | 131,758 | 2.28 | 1 | 132,707 | 0.75 |
|  | **36-45** | 4 | 144,394 | 2.77 | 8 | 147,561 | 5.42 | 2 | 149,845 | 1.33 | 3 | 150,593 | 1.99 |
|  | **46-55** | 4 | 158,162 | 2.53 | 4 | 161,272 | 2.48 | 7 | 163,200 | 4.29 | 7 | 163,550 | 4.28 |
|  | **56-65** | 10 | 142,998 | 6.99 | 7 | 144,059 | 4.86 | 6 | 144,191 | 4.16 | 12 | 143,017 | 8.39 |
|  | **Total** | 19 | 572,154 | 3.32 | 19 | 582,270 | 3.26 | 18 | 588,995 | 3.06 | 23 | 589,867 | 3.9 |
| **Total** | **26-35** | 4 | 248,297 | 1.61 | 7 | 253,266 | 2.76 | 10 | 257,148 | 3.89 | 6 | 259,305 | 2.31 |
|  | **36-45** | 8 | 284,838 | 2.81 | 19 | 290,710 | 6.54 | 11 | 294,616 | 3.73 | 16 | 296,347 | 5.4 |
|  | **46-55** | 20 | 315,870 | 6.33 | 16 | 321,617 | 4.97 | 19 | 325,077 | 5.84 | 18 | 326,101 | 5.52 |
|  | **56-65** | 13 | 288,974 | 4.5 | 17 | 290,818 | 5.85 | 11 | 290,452 | 3.79 | 33 | 287,989 | 11.46 |
|  | **Total** | 45 | 1,137,979 | 3.95 | 59 | 1,156,412 | 5.1 | 51 | 1,167,292 | 4.37 | 73 | 1,169,742 | 6.24 |
| CASE is CVT with or without other diagnoses | | | | |  |  |  |  |  |  |  |  |  |

**QUARTERLY INCIDENCE RATES: CVT 2016 (**Cases per 100,000 Person-Years)

|  |  | **2016 Q1** | | | **2016 Q2** | | | **2016 Q3** | | | **2016 Q4** | | |
| --- | --- | --- | --- | --- | --- | --- | --- | --- | --- | --- | --- | --- | --- |
|  | **Age Group** | **Cases** | **Person-Years** | **Incidence** | **Cases** | **Person-Years** | **Incidence** | **Cases** | **Person-Years** | **Incidence** | **Cases** | **Person-Years** | **Incidence** |
| **Female** | **26-35** | 13 | 127,613 | 10.19 | 10 | 131,904 | 7.58 | 9 | 135,676 | 6.63 | 13 | 139,619 | 9.31 |
|  | **36-45** | 14 | 146,554 | 9.55 | 13 | 150,480 | 8.64 | 18 | 153,921 | 11.69 | 11 | 157,492 | 6.98 |
|  | **46-55** | 9 | 165,941 | 5.42 | 12 | 169,697 | 7.07 | 17 | 172,730 | 9.84 | 14 | 175,604 | 7.97 |
|  | **56-65** | 16 | 160,290 | 9.98 | 13 | 162,519 | 8.00 | 10 | 163,186 | 6.13 | 19 | 163,806 | 11.6 |
|  | **Total** | 52 | 600,398 | 8.66 | 48 | 614,600 | 7.81 | 54 | 625,512 | 8.63 | 57 | 636,521 | 8.95 |
| **Male** | **26-35** | 2 | 135,289 | 1.48 | 2 | 141,286 | 1.42 | 6 | 146,172 | 4.10 | 7 | 149,789 | 4.67 |
|  | **36-45** | 5 | 151,197 | 3.31 | 4 | 156,829 | 2.55 | 6 | 161,086 | 3.72 | 2 | 164,508 | 1.22 |
|  | **46-55** | 5 | 166,343 | 3.01 | 6 | 170,845 | 3.51 | 13 | 174,243 | 7.46 | 8 | 177,097 | 4.52 |
|  | **56-65** | 17 | 155,880 | 10.91 | 16 | 158,178 | 10.12 | 6 | 159,030 | 3.77 | 12 | 159,808 | 7.51 |
|  | **Total** | 29 | 608,709 | 4.76 | 28 | 627,138 | 4.46 | 31 | 640,531 | 4.84 | 29 | 651,203 | 4.45 |
| **Total** | **26-35** | 15 | 262,902 | 5.71 | 12 | 273,190 | 4.39 | 15 | 281,848 | 5.32 | 20 | 289,408 | 6.91 |
|  | **36-45** | 19 | 297,751 | 6.38 | 17 | 307,309 | 5.53 | 24 | 315,007 | 7.62 | 13 | 322,001 | 4.04 |
|  | **46-55** | 14 | 332,284 | 4.21 | 18 | 340,542 | 5.29 | 30 | 346,972 | 8.65 | 22 | 352,701 | 6.24 |
|  | **56-65** | 33 | 316,170 | 10.44 | 29 | 320,697 | 9.04 | 16 | 322,216 | 4.97 | 31 | 323,614 | 9.58 |
|  | **Total** | 81 | 1,209,106 | 6.70 | 76 | 1,241,738 | 6.12 | 85 | 1,266,043 | 6.71 | 86 | 1,287,724 | 6.68 |
| CASE is CVT with or without other diagnoses | | | | |  |  |  |  |  |  |  |  |  |

**QUARTERLY INCIDENCE RATES: CVT 2017 (**Cases per 100,000 Person-Years)

|  |  | **2017 Q1** | | | **2017 Q2** | | | **2017 Q3** | | | **2017 Q4** | | |
| --- | --- | --- | --- | --- | --- | --- | --- | --- | --- | --- | --- | --- | --- |
|  | **Age Group** | **Cases** | **Person-Years** | **Incidence** | **Cases** | **Person-Years** | **Incidence** | **Cases** | **Person-Years** | **Incidence** | **Cases** | **Person-Years** | **Incidence** |
| **Female** | **26-35** | 19 | 135,064 | 14.07 | 7 | 136,717 | 5.12 | 11 | 141,107 | 7.80 | 16 | 145,158 | 11.02 |
|  | **36-45** | 12 | 152,370 | 7.88 | 8 | 153,945 | 5.20 | 10 | 158,028 | 6.33 | 16 | 161,642 | 9.90 |
|  | **46-55** | 13 | 170,419 | 7.63 | 18 | 171,863 | 10.47 | 6 | 176,041 | 3.41 | 15 | 179,646 | 8.35 |
|  | **56-65** | 12 | 171,083 | 7.01 | 13 | 171,112 | 7.60 | 18 | 173,437 | 10.38 | 20 | 174,816 | 11.44 |
|  | **Total** | 56 | 628,935 | 8.90 | 46 | 633,638 | 7.26 | 45 | 648,613 | 6.94 | 67 | 661,262 | 10.13 |
| **Male** | **26-35** | 4 | 146,712 | 2.73 | 3 | 149,075 | 2.01 | 6 | 153,720 | 3.90 | 2 | 157,834 | 1.27 |
|  | **36-45** | 6 | 160,287 | 3.74 | 6 | 162,714 | 3.69 | 4 | 167,399 | 2.39 | 5 | 171,410 | 2.92 |
|  | **46-55** | 9 | 173,222 | 5.20 | 5 | 175,095 | 2.86 | 7 | 179,560 | 3.90 | 5 | 183,484 | 2.73 |
|  | **56-65** | 16 | 167,592 | 9.55 | 11 | 168,045 | 6.55 | 10 | 170,665 | 5.86 | 11 | 172,423 | 6.38 |
|  | **Total** | 35 | 647,812 | 5.40 | 25 | 654,930 | 3.82 | 27 | 671,344 | 4.02 | 23 | 685,152 | 3.36 |
| **Total** | **26-35** | 23 | 281,775 | 8.16 | 10 | 285,793 | 3.50 | 17 | 294,827 | 5.77 | 18 | 302,992 | 5.94 |
|  | **36-45** | 18 | 312,656 | 5.76 | 14 | 316,659 | 4.42 | 14 | 325,427 | 4.30 | 21 | 333,052 | 6.31 |
|  | **46-55** | 22 | 343,641 | 6.40 | 23 | 346,959 | 6.63 | 13 | 355,601 | 3.66 | 20 | 363,130 | 5.51 |
|  | **56-65** | 28 | 338,675 | 8.27 | 24 | 339,157 | 7.08 | 28 | 344,102 | 8.14 | 31 | 347,239 | 8.93 |
|  | **Total** | 91 | 1,276,747 | 7.13 | 71 | 1,288,567 | 5.51 | 72 | 1,319,957 | 5.45 | 90 | 1,346,414 | 6.68 |
| CASE is CVT with or without other diagnoses | | | | |  |  |  |  |  |  |  |  |  |

**QUARTERLY INCIDENCE RATES: CVT 2018 (**Cases per 100,000 Person-Years)

|  |  | **2018 Q1** | | | **2018 Q2** | | | **2018 Q3** | | | **2018 Q4** | | |
| --- | --- | --- | --- | --- | --- | --- | --- | --- | --- | --- | --- | --- | --- |
|  | **Age Group** | **Cases** | **Person-Years** | **Incidence** | **Cases** | **Person-Years** | **Incidence** | **Cases** | **Person-Years** | **Incidence** | **Cases** | **Person-Years** | **Incidence** |
| **Female** | **26-35** | 9 | 142,779 | 6.3 | 4 | 145,153 | 2.76 | 14 | 146,789 | 9.54 | 18 | 148,051 | 12.16 |
|  | **36-45** | 13 | 160,729 | 8.09 | 21 | 163,660 | 12.83 | 16 | 165,707 | 9.66 | 20 | 166,873 | 11.99 |
|  | **46-55** | 18 | 175,504 | 10.26 | 15 | 178,471 | 8.4 | 20 | 180,720 | 11.07 | 15 | 181,897 | 8.25 |
|  | **56-65** | 10 | 182,823 | 5.47 | 11 | 184,459 | 5.96 | 19 | 185,402 | 10.25 | 17 | 185,093 | 9.18 |
|  | **Total** | 50 | 661,835 | 7.55 | 51 | 671,743 | 7.59 | 69 | 678,619 | 10.17 | 70 | 681,915 | 10.27 |
| **Male** | **26-35** | 3 | 156,820 | 1.91 | 4 | 159,226 | 2.51 | 4 | 161,214 | 2.48 | 8 | 162,558 | 4.92 |
|  | **36-45** | 6 | 171,974 | 3.49 | 6 | 174,737 | 3.43 | 4 | 176,980 | 2.26 | 9 | 178,058 | 5.05 |
|  | **46-55** | 11 | 181,113 | 6.07 | 8 | 183,900 | 4.35 | 9 | 186,294 | 4.83 | 14 | 187,380 | 7.47 |
|  | **56-65** | 16 | 181,477 | 8.82 | 12 | 182,998 | 6.56 | 15 | 184,056 | 8.15 | 14 | 183,751 | 7.62 |
|  | **Total** | 36 | 691,384 | 5.21 | 30 | 700,860 | 4.28 | 32 | 708,545 | 4.52 | 45 | 711,748 | 6.32 |
| **Total** | **26-35** | 12 | 299,598 | 4.01 | 8 | 304,379 | 2.63 | 18 | 308,004 | 5.84 | 26 | 310,609 | 8.37 |
|  | **36-45** | 19 | 332,704 | 5.71 | 27 | 338,397 | 7.98 | 20 | 342,687 | 5.84 | 29 | 344,931 | 8.41 |
|  | **46-55** | 29 | 356,617 | 8.13 | 23 | 362,370 | 6.35 | 29 | 367,014 | 7.9 | 29 | 369,277 | 7.85 |
|  | **56-65** | 26 | 364,300 | 7.14 | 23 | 367,457 | 6.26 | 34 | 369,458 | 9.2 | 31 | 368,844 | 8.4 |
|  | **Total** | 86 | 1,353,219 | 6.36 | 81 | 1,372,603 | 5.9 | 101 | 1,387,163 | 7.28 | 115 | 1,393,662 | 8.25 |
| CASE is CVT with or without other diagnoses | | | | |  |  |  |  |  |  |  |  |  |

**QUARTERLY INCIDENCE RATES: CVT 2019 (**Cases per 100,000 Person-Years)

|  |  | **2019 Q1** | | | **2019 Q2** | | | **2019 Q3** | | | **2019 Q4** | | |
| --- | --- | --- | --- | --- | --- | --- | --- | --- | --- | --- | --- | --- | --- |
|  | **Age Group** | **Cases** | **Person-Years** | **Incidence** | **Cases** | **Person-Years** | **Incidence** | **Cases** | **Person-Years** | **Incidence** | **Cases** | **Person-Years** | **Incidence** |
| **Female** | **26-35** | 12 | 143,302 | 8.37 | 18 | 145,542 | 12.37 | 23 | 147,626 | 15.58 | 20 | 149,168 | 13.41 |
|  | **36-45** | 9 | 162,877 | 5.53 | 27 | 165,344 | 16.33 | 19 | 167,434 | 11.35 | 20 | 168,212 | 11.89 |
|  | **46-55** | 20 | 174,682 | 11.45 | 19 | 177,040 | 10.73 | 10 | 179,085 | 5.58 | 21 | 179,889 | 11.67 |
|  | **56-65** | 12 | 188,366 | 6.37 | 20 | 189,422 | 10.56 | 13 | 190,433 | 6.83 | 21 | 189,828 | 11.06 |
|  | **Total** | 53 | 669,227 | 7.92 | 84 | 677,348 | 12.4 | 65 | 684,578 | 9.49 | 82 | 687,097 | 11.93 |
| **Male** | **26-35** | 4 | 157,453 | 2.54 | 3 | 159,571 | 1.88 | 3 | 161,210 | 1.86 | 4 | 162,255 | 2.47 |
|  | **36-45** | 5 | 173,901 | 2.88 | 1 | 176,149 | 0.57 | 8 | 177,670 | 4.5 | 1 | 178,207 | 0.56 |
|  | **46-55** | 12 | 180,223 | 6.66 | 10 | 182,497 | 5.48 | 9 | 184,349 | 4.88 | 10 | 184,933 | 5.41 |
|  | **56-65** | 21 | 186,104 | 11.28 | 18 | 187,160 | 9.62 | 13 | 188,161 | 6.91 | 19 | 187,566 | 10.13 |
|  | **Total** | 42 | 697,681 | 6.02 | 32 | 705,378 | 4.54 | 33 | 711,389 | 4.64 | 34 | 712,962 | 4.77 |
| **Total** | **26-35** | 16 | 300,756 | 5.32 | 21 | 305,113 | 6.88 | 26 | 308,836 | 8.42 | 24 | 311,423 | 7.71 |
|  | **36-45** | 14 | 336,778 | 4.16 | 28 | 341,493 | 8.2 | 27 | 345,104 | 7.82 | 21 | 346,419 | 6.06 |
|  | **46-55** | 32 | 354,905 | 9.02 | 29 | 359,537 | 8.07 | 19 | 363,433 | 5.23 | 31 | 364,822 | 8.50 |
|  | **56-65** | 33 | 374,470 | 8.81 | 38 | 376,582 | 10.09 | 26 | 378,595 | 6.87 | 40 | 377,395 | 10.6 |
|  | **Total** | 95 | 1,366,908 | 6.95 | 116 | 1,382,726 | 8.39 | 98 | 1,395,968 | 7.02 | 116 | 1,400,059 | 8.29 |

CASE is CVT with or without other diagnoses

**QUARTERLY INCIDENCE RATES: CVT first quarter 2020 (**Cases per 100,000 Person-Years)

|  |  | **2020 Q1** | | |
| --- | --- | --- | --- | --- |
|  | **Age Group** | **Cases** | **Person-Years** | **Incidence** |
| **Female** | **26-35** | 12 | 139,751 | 8.59 |
|  | **36-45** | 20 | 159,276 | 12.56 |
|  | **46-55** | 19 | 168,194 | 11.30 |
|  | **56-65** | 19 | 188,937 | 10.06 |
|  | **Total** | 70 | 656,158 | 10.67 |
| **Male** | **26-35** | 0 | 152,223 | 0 |
|  | **36-45** | 9 | 169,385 | 5.31 |
|  | **46-55** | 14 | 173,844 | 8.05 |
|  | **56-65** | 22 | 186,497 | 11.8 |
|  | **Total** | 45 | 681,949 | 6.60 |
| **Total** | **26-35** | 12 | 291,974 | 4.11 |
|  | **36-45** | 29 | 328,662 | 8.82 |
|  | **46-55** | 33 | 342,037 | 9.65 |
|  | **56-65** | 41 | 375,434 | 10.92 |
|  | **Total** | 115 | 1,338,107 | 8.59 |
| CASE is CVT with or without other diagnoses | | | | |

**QUARTERLY INCIDENCE RATES: ANY MAJOR VENOUS THROMBOSIS 2015 (Cases per 100,000 Person-Years)**

|  |  | **2015 Q1** | | | **2015 Q2** | | | **2015 Q3** | | | **2015 Q4** | | |
| --- | --- | --- | --- | --- | --- | --- | --- | --- | --- | --- | --- | --- | --- |
|  | **Age Group** | **Cases** | **Person-Years** | **Incidence** | **Cases** | **Person-Years** | **Incidence** | **Cases** | **Person-Years** | **Incidence** | **Cases** | **Person-Years** | **Incidence** |
| **Female** | **26-35** | 179 | 121,470 | 147.36 | 181 | 123,650 | 146.38 | 189 | 125,149 | 151.02 | 147 | 126,354 | 116.34 |
|  | **36-45** | 299 | 139,978 | 213.6 | 268 | 142,678 | 187.84 | 341 | 144,305 | 236.3 | 279 | 145,281 | 192.04 |
|  | **46-55** | 467 | 156,926 | 297.59 | 486 | 159,544 | 304.62 | 501 | 161,071 | 311.04 | 457 | 161,745 | 282.54 |
|  | **56-65** | 693 | 144,849 | 478.43 | 695 | 145,612 | 477.29 | 700 | 145,129 | 482.33 | 662 | 143,852 | 460.19 |
|  | **Total** | 1,638 | 563,223 | 290.83 | 1,630 | 571,484 | 285.22 | 1,731 | 575,655 | 300.7 | 1,545 | 577,232 | 267.66 |
| **Male** | **26-35** | 86 | 126,468 | 68 | 86 | 129,242 | 66.54 | 95 | 131,619 | 72.18 | 71 | 132,567 | 53.56 |
|  | **36-45** | 215 | 144,056 | 149.25 | 206 | 147,216 | 139.93 | 222 | 149,493 | 148.5 | 202 | 150,236 | 134.45 |
|  | **46-55** | 486 | 157,383 | 308.8 | 446 | 160,479 | 277.92 | 503 | 162,394 | 309.74 | 456 | 162,731 | 280.22 |
|  | **56-65** | 755 | 141,683 | 532.88 | 801 | 142,728 | 561.21 | 789 | 142,866 | 552.26 | 774 | 141,703 | 546.21 |
|  | **Total** | 1,542 | 569,590 | 270.72 | 1,539 | 579,665 | 265.5 | 1,609 | 586,373 | 274.4 | 1,503 | 587,238 | 255.94 |
| **Total** | **26-35** | 265 | 247,938 | 106.88 | 267 | 252,892 | 105.58 | 284 | 256,768 | 110.61 | 218 | 258,922 | 84.2 |
|  | **36-45** | 514 | 284,035 | 180.96 | 474 | 289,895 | 163.51 | 563 | 293,799 | 191.63 | 481 | 295,517 | 162.77 |
|  | **46-55** | 953 | 314,309 | 303.21 | 932 | 320,023 | 291.23 | 1,004 | 323,466 | 310.39 | 913 | 324,477 | 281.38 |
|  | **56-65** | 1,448 | 286,531 | 505.35 | 1,496 | 288,340 | 518.83 | 1,489 | 287,995 | 517.02 | 1,436 | 285,556 | 502.88 |
|  | **Total** | 3,180 | 1,132,813 | 280.72 | 3,169 | 1,151,149 | 275.29 | 3,340 | 1,162,027 | 287.43 | 3,048 | 1,164,471 | 261.75 |
| Any of the above (DVT, PE, CVT or Any of the following: ) | | | | | |  |  |  |  |  |  |  |  |
| Portal vein thrombosis | |  |  |  |  |  |  |  |  |  |  |  |  |
| Hepatic vein thrombosis/Budd-Chiari syndrome | | | | |  |  |  |  |  |  |  |  |  |
| Thrombophlebitis migrans | | |  |  |  |  |  |  |  |  |  |  |  |
| Embolism or thrombosis of vena cava (inferior) | | | | |  |  |  |  |  |  |  |  |  |
| Embolism or thrombosis of renal vein | | | |  |  |  |  |  |  |  |  |  |  |
| Mesenteric thrombosis | | |  |  |  |  |  |  |  |  |  |  |  |

**QUARTERLY INCIDENCE RATES: ANY MAJOR VENOUS THROMBOSIS 2016 (Cases per 100,000 Person-Years)**

|  |  | **2016 Q1** | | | **2016 Q2** | | | **2016 Q3** | | | **2016 Q4** | | |
| --- | --- | --- | --- | --- | --- | --- | --- | --- | --- | --- | --- | --- | --- |
|  | **Age Group** | **Cases** | **Person-Years** | **Incidence** | **Cases** | **Person-Years** | **Incidence** | **Cases** | **Person-Years** | **Incidence** | **Cases** | **Person-Years** | **Incidence** |
| **Female** | **26-35** | 131 | 127,388 | 102.84 | 133 | 131,686 | 101 | 176 | 135,462 | 129.93 | 158 | 139,409 | 113.34 |
|  | **36-45** | 301 | 146,092 | 206.03 | 271 | 150,014 | 180.65 | 308 | 153,453 | 200.71 | 306 | 157,033 | 194.86 |
|  | **46-55** | 446 | 165,144 | 270.07 | 448 | 168,901 | 265.24 | 501 | 171,941 | 291.38 | 488 | 174,830 | 279.13 |
|  | **56-65** | 700 | 159,128 | 439.9 | 679 | 161,357 | 420.8 | 662 | 162,051 | 408.51 | 669 | 162,727 | 411.12 |
|  | **Total** | 1,578 | 597,752 | 263.99 | 1,531 | 611,958 | 250.18 | 1,647 | 622,907 | 264.41 | 1,621 | 633,999 | 255.68 |
| **Male** | **26-35** | 62 | 135,160 | 45.87 | 96 | 141,156 | 68.01 | 83 | 146,041 | 56.83 | 99 | 149,663 | 66.15 |
|  | **36-45** | 207 | 150,865 | 137.21 | 218 | 156,506 | 139.29 | 204 | 160,757 | 126.9 | 218 | 164,178 | 132.78 |
|  | **46-55** | 423 | 165,570 | 255.48 | 434 | 170,065 | 255.2 | 472 | 173,462 | 272.11 | 486 | 176,331 | 275.62 |
|  | **56-65** | 810 | 154,496 | 524.29 | 799 | 156,779 | 509.64 | 770 | 157,658 | 488.4 | 804 | 158,476 | 507.33 |
|  | **Total** | 1,502 | 606,092 | 247.82 | 1,547 | 624,505 | 247.72 | 1,529 | 637,918 | 239.69 | 1,607 | 648,647 | 247.75 |
| **Total** | **26-35** | 193 | 262,548 | 73.51 | 229 | 272,842 | 83.93 | 259 | 281,503 | 92.01 | 257 | 289,072 | 88.91 |
|  | **36-45** | 508 | 296,957 | 171.07 | 489 | 306,519 | 159.53 | 512 | 314,209 | 162.95 | 524 | 321,211 | 163.13 |
|  | **46-55** | 869 | 330,715 | 262.76 | 882 | 338,966 | 260.2 | 973 | 345,403 | 281.7 | 974 | 351,161 | 277.37 |
|  | **56-65** | 1,510 | 313,624 | 481.47 | 1,478 | 318,136 | 464.58 | 1,432 | 319,709 | 447.91 | 1,473 | 321,202 | 458.59 |
|  | **Total** | 3,080 | 1,203,844 | 255.85 | 3,078 | 1,236,463 | 248.94 | 3,176 | 1,260,825 | 251.9 | 3,228 | 1,282,646 | 251.67 |
| Any of the above (DVT, PE, CVT or Any of the following: ) | | | | | |  |  |  |  |  |  |  |  |
| Portal vein thrombosis | |  |  |  |  |  |  |  |  |  |  |  |  |
| Hepatic vein thrombosis/Budd-Chiari syndrome | | | | |  |  |  |  |  |  |  |  |  |
| Thrombophlebitis migrans | | |  |  |  |  |  |  |  |  |  |  |  |
| Embolism or thrombosis of vena cava (inferior) | | | | |  |  |  |  |  |  |  |  |  |
| Embolism or thrombosis of renal vein | | | |  |  |  |  |  |  |  |  |  |  |
| Mesenteric thrombosis | | |  |  |  |  |  |  |  |  |  |  |  |

**QUARTERLY INCIDENCE RATES: ANY MAJOR VENOUS THROMBOSIS 2017 (Cases per 100,000 Person-Years)**

|  |  | **2017 Q1** | | | **2017 Q2** | | | **2017 Q3** | | | **2017 Q4** | | |
| --- | --- | --- | --- | --- | --- | --- | --- | --- | --- | --- | --- | --- | --- |
|  | **Age Group** | **Cases** | **Person-Years** | **Incidence** | **Cases** | **Person-Years** | **Incidence** | **Cases** | **Person-Years** | **Incidence** | **Cases** | **Person-Years** | **Incidence** |
| **Female** | **26-35** | 157 | 134,862 | 116.42 | 132 | 136,515 | 96.69 | 165 | 140,891 | 117.11 | 132 | 144,932 | 91.08 |
|  | **36-45** | 293 | 151,930 | 192.85 | 277 | 153,502 | 180.45 | 313 | 157,572 | 198.64 | 288 | 161,175 | 178.69 |
|  | **46-55** | 533 | 169,628 | 314.22 | 462 | 171,067 | 270.07 | 452 | 175,219 | 257.96 | 502 | 178,818 | 280.73 |
|  | **56-65** | 786 | 169,913 | 462.59 | 754 | 169,919 | 443.74 | 803 | 172,210 | 466.29 | 837 | 173,555 | 482.27 |
|  | **Total** | 1,769 | 626,333 | 282.44 | 1,625 | 631,003 | 257.53 | 1,733 | 645,892 | 268.31 | 1,759 | 658,480 | 267.13 |
| **Male** | **26-35** | 75 | 146,597 | 51.16 | 96 | 148,954 | 64.45 | 93 | 153,593 | 60.55 | 95 | 157,701 | 60.24 |
|  | **36-45** | 225 | 159,974 | 140.65 | 220 | 162,394 | 135.47 | 207 | 167,065 | 123.9 | 204 | 171,061 | 119.26 |
|  | **46-55** | 460 | 172,492 | 266.68 | 474 | 174,353 | 271.86 | 477 | 178,790 | 266.79 | 482 | 182,692 | 263.83 |
|  | **56-65** | 900 | 166,199 | 541.52 | 844 | 166,651 | 506.45 | 883 | 169,249 | 521.72 | 896 | 170,981 | 524.03 |
|  | **Total** | 1,660 | 645,262 | 257.26 | 1,634 | 652,352 | 250.48 | 1,660 | 668,697 | 248.24 | 1,677 | 682,435 | 245.74 |
| **Total** | **26-35** | 232 | 281,460 | 82.43 | 228 | 285,469 | 79.87 | 258 | 294,483 | 87.61 | 227 | 302,633 | 75.01 |
|  | **36-45** | 518 | 311,904 | 166.08 | 497 | 315,895 | 157.33 | 520 | 324,638 | 160.18 | 492 | 332,236 | 148.09 |
|  | **46-55** | 993 | 342,120 | 290.25 | 936 | 345,420 | 270.97 | 929 | 354,009 | 262.42 | 984 | 361,510 | 272.19 |
|  | **56-65** | 1,686 | 336,112 | 501.62 | 1,598 | 336,570 | 474.79 | 1,686 | 341,459 | 493.76 | 1,733 | 344,537 | 502.99 |
|  | **Total** | 3,429 | 1,271,595 | 269.66 | 3,259 | 1,283,355 | 253.94 | 3,393 | 1,314,589 | 258.1 | 3,436 | 1,340,916 | 256.24 |
| Any of the above (DVT, PE, CVT or Any of the following: ) | | | | | |  |  |  |  |  |  |  |  |
| Portal vein thrombosis | |  |  |  |  |  |  |  |  |  |  |  |  |
| Hepatic vein thrombosis/Budd-Chiari syndrome | | | | |  |  |  |  |  |  |  |  |  |
| Thrombophlebitis migrans | | |  |  |  |  |  |  |  |  |  |  |  |
| Embolism or thrombosis of vena cava (inferior) | | | | |  |  |  |  |  |  |  |  |  |
| Embolism or thrombosis of renal vein | | | |  |  |  |  |  |  |  |  |  |  |
| Mesenteric thrombosis | | |  |  |  |  |  |  |  |  |  |  |  |

**QUARTERLY INCIDENCE RATES: ANY MAJOR VENOUS THROMBOSIS 2018 (Cases per 100,000 Person-Years)**

|  |  | **2018 Q1** | | | **2018 Q2** | | | **2018 Q3** | | | **2018 Q4** | | |
| --- | --- | --- | --- | --- | --- | --- | --- | --- | --- | --- | --- | --- | --- |
|  | **Age Group** | **Cases** | **Person-Years** | **Incidence** | **Cases** | **Person-Years** | **Incidence** | **Cases** | **Person-Years** | **Incidence** | **Cases** | **Person-Years** | **Incidence** |
| **Female** | **26-35** | 129 | 142,566 | 90.48 | 135 | 144,942 | 93.14 | 162 | 146,574 | 110.52 | 172 | 147,826 | 116.35 |
|  | **36-45** | 309 | 160,295 | 192.77 | 308 | 163,210 | 188.71 | 319 | 165,253 | 193.04 | 338 | 166,400 | 203.12 |
|  | **46-55** | 517 | 174,745 | 295.86 | 495 | 177,687 | 278.58 | 548 | 179,917 | 304.58 | 550 | 181,072 | 303.75 |
|  | **56-65** | 852 | 181,528 | 469.35 | 861 | 183,130 | 470.16 | 839 | 184,049 | 455.86 | 793 | 183,731 | 431.61 |
|  | **Total** | 1,807 | 659,134 | 274.15 | 1,799 | 668,968 | 268.92 | 1,868 | 675,793 | 276.42 | 1,853 | 679,029 | 272.89 |
| **Male** | **26-35** | 84 | 156,699 | 53.61 | 104 | 159,099 | 65.37 | 94 | 161,075 | 58.36 | 97 | 162,414 | 59.72 |
|  | **36-45** | 189 | 171,656 | 110.1 | 190 | 174,415 | 108.94 | 221 | 176,655 | 125.1 | 215 | 177,725 | 120.97 |
|  | **46-55** | 513 | 180,369 | 284.42 | 496 | 183,142 | 270.83 | 512 | 185,524 | 275.98 | 505 | 186,587 | 270.65 |
|  | **56-65** | 950 | 179,994 | 527.79 | 965 | 181,464 | 531.79 | 978 | 182,498 | 535.9 | 1,012 | 182,169 | 555.53 |
|  | **Total** | 1,736 | 688,719 | 252.06 | 1,755 | 698,119 | 251.39 | 1,805 | 705,752 | 255.76 | 1,829 | 708,896 | 258.01 |
| **Total** | **26-35** | 213 | 299,265 | 71.17 | 239 | 304,040 | 78.61 | 256 | 307,649 | 83.21 | 269 | 310,240 | 86.71 |
|  | **36-45** | 498 | 331,951 | 150.02 | 498 | 337,625 | 147.5 | 540 | 341,909 | 157.94 | 553 | 344,125 | 160.7 |
|  | **46-55** | 1,030 | 355,114 | 290.05 | 991 | 360,828 | 274.65 | 1,060 | 365,441 | 290.06 | 1,055 | 367,659 | 286.95 |
|  | **56-65** | 1,802 | 361,523 | 498.45 | 1,826 | 364,594 | 500.83 | 1,817 | 366,547 | 495.71 | 1,805 | 365,900 | 493.3 |
|  | **Total** | 3,543 | 1,347,852 | 262.86 | 3,554 | 1,367,087 | 259.97 | 3,673 | 1,381,545 | 265.86 | 3,682 | 1,387,925 | 265.29 |
| Any of the above (DVT, PE, CVT or Any of the following: ) | | | | | |  |  |  |  |  |  |  |  |
| Portal vein thrombosis | |  |  |  |  |  |  |  |  |  |  |  |  |
| Hepatic vein thrombosis/Budd-Chiari syndrome | | | | |  |  |  |  |  |  |  |  |  |
| Thrombophlebitis migrans | | |  |  |  |  |  |  |  |  |  |  |  |
| Embolism or thrombosis of vena cava (inferior) | | | | |  |  |  |  |  |  |  |  |  |
| Embolism or thrombosis of renal vein | | | |  |  |  |  |  |  |  |  |  |  |
| Mesenteric thrombosis | | |  |  |  |  |  |  |  |  |  |  |  |

**QUARTERLY INCIDENCE RATES: ANY MAJOR VENOUS THROMBOSIS 2019 (Cases per 100,000 Person-Years)**

|  |  | **2019 Q1** | | | **2019 Q2** | | | **2019 Q3** | | | **2019 Q4** | | |
| --- | --- | --- | --- | --- | --- | --- | --- | --- | --- | --- | --- | --- | --- |
|  | **Age Group** | **Cases** | **Person-Years** | **Incidence** | **Cases** | **Person-Years** | **Incidence** | **Cases** | **Person-Years** | **Incidence** | **Cases** | **Person-Years** | **Incidence** |
| **Female** | **26-35** | 156 | 143,091 | 109.02 | 170 | 145,323 | 116.98 | 170 | 147,389 | 115.34 | 172 | 148,928 | 115.49 |
|  | **36-45** | 311 | 162,432 | 191.47 | 337 | 164,888 | 204.38 | 328 | 166,957 | 196.46 | 306 | 167,724 | 182.44 |
|  | **46-55** | 536 | 173,892 | 308.24 | 547 | 176,210 | 310.42 | 535 | 178,225 | 300.18 | 514 | 179,005 | 287.14 |
|  | **56-65** | 891 | 187,027 | 476.4 | 874 | 188,057 | 464.75 | 887 | 189,032 | 469.23 | 886 | 188,403 | 470.27 |
|  | **Total** | 1,894 | 666,442 | 284.2 | 1,928 | 674,478 | 285.85 | 1,920 | 681,603 | 281.69 | 1,878 | 684,060 | 274.54 |
| **Male** | **26-35** | 91 | 157,330 | 57.84 | 100 | 159,441 | 62.72 | 106 | 161,072 | 65.81 | 100 | 162,115 | 61.68 |
|  | **36-45** | 192 | 173,603 | 110.6 | 210 | 175,836 | 119.43 | 233 | 177,341 | 131.39 | 228 | 177,865 | 128.19 |
|  | **46-55** | 498 | 179,499 | 277.44 | 469 | 181,749 | 258.05 | 480 | 183,583 | 261.46 | 491 | 184,160 | 266.62 |
|  | **56-65** | 998 | 184,550 | 540.78 | 983 | 185,596 | 529.64 | 985 | 186,571 | 527.95 | 1,044 | 185,953 | 561.43 |
|  | **Total** | 1,779 | 694,982 | 255.98 | 1,762 | 702,622 | 250.78 | 1,804 | 708,567 | 254.6 | 1,863 | 710,093 | 262.36 |
| **Total** | **26-35** | 247 | 300,421 | 82.22 | 270 | 304,764 | 88.59 | 276 | 308,461 | 89.48 | 272 | 311,043 | 87.45 |
|  | **36-45** | 503 | 336,035 | 149.69 | 547 | 340,724 | 160.54 | 561 | 344,298 | 162.94 | 534 | 345,589 | 154.52 |
|  | **46-55** | 1,034 | 353,391 | 292.59 | 1,016 | 357,959 | 283.83 | 1,015 | 361,808 | 280.54 | 1,005 | 363,165 | 276.73 |
|  | **56-65** | 1,889 | 371,577 | 508.37 | 1,857 | 373,653 | 496.99 | 1,872 | 375,603 | 498.4 | 1,930 | 374,356 | 515.55 |
|  | **Total** | 3,673 | 1,361,424 | 269.79 | 3,690 | 1,377,100 | 267.95 | 3,724 | 1,390,170 | 267.88 | 3,741 | 1,394,153 | 268.33 |
| Any of the above (DVT, PE, CVT or Any of the following: ) | | | | | |  |  |  |  |  |  |  |  |
| Portal vein thrombosis | |  |  |  |  |  |  |  |  |  |  |  |  |
| Hepatic vein thrombosis/Budd-Chiari syndrome | | | | |  |  |  |  |  |  |  |  |  |
| Thrombophlebitis migrans | | |  |  |  |  |  |  |  |  |  |  |  |
| Embolism or thrombosis of vena cava (inferior) | | | | |  |  |  |  |  |  |  |  |  |
| Embolism or thrombosis of renal vein | | | |  |  |  |  |  |  |  |  |  |  |
| Mesenteric thrombosis | | |  |  |  |  |  |  |  |  |  |  |  |

**QUARTERLY INCIDENCE RATES: ANY MAJOR VENOUS THROMBOSIS first quarter 2020 (**Cases per 100,000 Person-Years)

|  |  | **2020 Q1** | | |
| --- | --- | --- | --- | --- |
|  | **Age Group** | **Cases** | **Person-Years** | **Incidence** |
| **Female** | **26-35** | 139 | 139,540 | 99.61 |
|  | **36-45** | 293 | 158,835 | 184.47 |
|  | **46-55** | 487 | 167,375 | 290.96 |
|  | **56-65** | 839 | 187,540 | 447.37 |
|  | **Total** | 1,758 | 653,291 | 269.1 |
| **Male** | **26-35** | 91 | 152,095 | 59.83 |
|  | **36-45** | 206 | 169,064 | 121.85 |
|  | **46-55** | 501 | 173,129 | 289.38 |
|  | **56-65** | 1,005 | 184,926 | 543.46 |
|  | **Total** | 1,803 | 679,215 | 265.45 |
| **Total** | **26-35** | 230 | 291,636 | 78.87 |
|  | **36-45** | 499 | 327,899 | 152.18 |
|  | **46-55** | 988 | 340,504 | 290.16 |
|  | **56-65** | 1,844 | 372,467 | 495.08 |
|  | **Total** | 3,561 | 1,332,505 | 267.24 |
| Any of the above (DVT, PE, CVT or Any of the following: ) | | | | |
| Portal vein thrombosis | |  |  |  |
| Hepatic vein thrombosis/Budd-Chiari syndrome | | | | |
| Thrombophlebitis migrans | | |  |  |
| Embolism or thrombosis of vena cava (inferior) | | | | |
| Embolism or thrombosis of renal vein | | | |  |
| Mesenteric thrombosis | | |  |  |

**Incidence SUMMARY with FIRST Quarter 2020**

| **Ranges for Quarterly Incidence Values 2014-2015 and First Quarter 2020** | | | | | | | | | |
| --- | --- | --- | --- | --- | --- | --- | --- | --- | --- |
| **Sub-Groups** | **N** | **2014-19 Mean** | **2014-19 Std Dev** | **2014-19 Median.** | **2014-19 Minimum** | **2014-19 Maximum** | **2019 Minimum** | **2019 Maximum** | **2020**  **First Qtr** |
| **DVT Females** | 20 | 15.427 | 5.60315 | 14.59 | 8.42 | 27.5 | 8.42 | 10.33 | 9.60 |
| **DVT Males** | 20 | 19.2095 | 6.48737 | 16.9 | 11.36 | 32.65 | 11.36 | 12.61 | 10.12 |
| **PE Females** | 20 | 118.0615 | 11.96183 | 119.255 | 96.13 | 134.72 | 129.57 | 133.43 | 131.93 |
| **PE Males** | 20 | 124.007 | 8.6527 | 121.51 | 103.58 | 143.49 | 119.45 | 143.49 | 145.75 |
| **VTE Females** | 20 | 131.8085 | 8.99791 | 134.09 | 115.42 | 144.44 | 137.72 | 143.65 | 141.56 |
| **VTE Males** | 20 | 141.3735 | 6.59697 | 140.735 | 131.12 | 154.19 | 131.12 | 154.19 | 156.21 |
| **CVT Females** | 20 | 8.525 | 1.90277 | 8.625 | 4.60 | 12.40 | 7.92 | 12.4 | 10.67 |
| **CVT Males** | 20 | 4.4475 | 0.87563 | 4.49 | 3.06 | 6.32 | 4.54 | 6.02 | 6.60 |

**SYSTEMATIC REVIEW TABLES FOR INCIDENCE ESTIMATES ACROSS STUDIES**

| **VTE INCIDENCE ESTIMATES – ALL STUDIES** | | | | | |
| --- | --- | --- | --- | --- | --- |
| **Author**  **[date]** | **Design** | **Outcomes** | **Incidence VTE**  **Per 100,000** | **Incidence DVT** | **Incidence PE ±DVT** |
| Silverstein [1998]^1^ | Mayo Health Care, MN cases compared to county population  Age: All  Time: 1966-1990  [1960-70-80-90 pop] [N=106,470 1990] | Review medical records first episode DVT or PE  [n=2214 15+ yrs] | Age- sex- adjust [census]:  149 [CI: 143-155] 15+ yrs  Males: 165/100,000  Female: 140/100,000 | Age-sex-adjust:  61 [CI: 57-65]  15+ yrs | Age-sex- adjust:  88 [CI: 83-92]  15+ yrs |
| Oger  [2002]^2^ | Brest Dist, France  3 Hospital centers  Age: all  Time: 1yr 1998-1999  (N=342,000) | Signs/Symptoms and/or imaging lower DVT or PE | 183 [CI: 169-198] | 124 [112-136] | 60 [CI:52-69] |
| Cushman [2004]^3^ | 2 US cohorts  [6 communities]  Age: 45+ years  Time: 7.6yr [1987-97]  [N=21,680] | Chart Review, ICD-9  Dx + imaging  [n=265 first VTE]  [n=39 recurrent VTE] | Crude:  161 [143-181] 45+ yrs Age- adjusted: 192  No sex diff for age <75 | Crude: 117 | Crude: 45 |
| White  [2005]^4^ | Calif Pat Discharge Data  Age: 18+ yrs  Time: 1 yr [1996]  [N= 23,343,083] | ICD-9-CM  Lower extremity DVT, PE, ante-partum VT & OB PE  n=21,002 new cases  [n=12,455 definite,  n=3,539 probable] | Crude: 69 [68-70] [def+prob cases]  Age- sex- race-adjusted:  Female: 93 [CI:92-95]  Male: 85 [CI: 84-87]  White: 103 [CI:101-105]  Sig dif by race/ethnicity | -- | -- |
| Naes  [2007]^5^ | County in Norway,  Hospital cases compared to county population  Age: 20+ yrs  Time: 1995-2001  [N=94,769] | Hospital discharge ICD -9 & 10  & imaging  [n=740 cases] | Crude: 143 [CI: 133-154]  standardized to world pop:  94 [86-101]  Female: 1.58/1,000  Male: 1.28/1,000  [no sex diff after age-adj] | Crude:  93 [CI: 85-102]  Stan world pop:  61 [CI: 55-67] | Crude:  50 [CI: 44-56]  Stan world pop  33 [CI: 28-37] |
| Huang  [2014]^6^ | Worcester, Mass  MSA residents  2 patient cohorts  Age: All  Duration: 1985-2009  [1985 N=379,953]  [2000 N=477,598] | Medical record review;  In- & out-patients  [n=3,887 first event]  [n=1138 recurrent] | Age- sex- adjust [census]:  108 [CI: 98-118] first event  [34 [CI: 28-40] recurrent]  Sig. increase over 25 yrs | Age-sex-adjust:  66 [CI: 59-74] first event  recurrent  25 [CI: 20-29] | Age-sex- adjust:  41 [CI: 35-47] first event  recurrent:  9.6 [6.6-12.6] |
| Payne [2021]^7^ | Meta-analysis  3 Provinces/studies, Canada  All cases, universal health care system  Age: All  Time: 2000-2012 | Cases determined by 1^st^ & 2^nd^ discharge diagnosis [IDC-9+10] & procedures | Meta-analysis: 129 [106-153]  Higher with older age |  |  |
| Delluc  [2016]^8^ | Brest Dist, France  4 Hospitals & network  Age: all  Time: 1yr 2013-2014  [N=367,911] | Chart review & imaging for symptomatic lower DVT or PE  [n=576] | 157 [CI: 144-169]  39 20–39 yrs  768 75+ yrs | 76 [CI: 67-85] | 81 [CI:72-90] |
| Ostergaard [2021]^9^ | Denmark, All cases, universal health care  National registry  Age: 18-99 yrs  Time: 2010-2018  N=4,915,426  Age: 18-64 yrs  [N= 3,963,153] | 1^st^ or 2^nd^ inpatient or out-pat diagnosis [ICD-10]  DVT, PE, & other venous thromboses. | For DVT+PE only:  170 [CI: 168-171] 18-99 yrs  91 [CI: 89-92] 18-64 yrs  VTE + other thromboses:  176 [CI: 175-178] 18-99yr  95 [CI: 94-96] 18-64 yrs  No sex diffs | -- | -- |
| Williame,  European database^10^ | Claims data from across Europe | VTE | Spain (BIFAP):  2017: 190.9 [CI: 188.1-193.7]  2018: 175.0 [CI: 172.4-177.7]  2019: 171.0 [CI: 168.3-173.3]  Spain (SIDAP):  2017: 182.5 [CI: 179.0-186.1]  2018: 190.0 [CI: 186.1-193.1]  2019: 200.2 [CI: 196.5-204.1]  Italy:  2017: 183.0 [CI: 178.2-187.9]  2018: 169.7 [CI: 165.2-174.4]  2019: 171.1 [CI: 166.5-175.7]  United Kingdom:  2017: 174.7 [CI: 170.5-179.1]  2018: 166.1 [CI: 161.9-170.5]  2019: 150.0 [CI: 145.8-154.2] | Spain:  98 [CI: 97-99]  Italy:  112 [109-114]  UK:  127 [125-129] | Spain:  55 [CI: 53,56]  Italy:  85 [CI: 83-86] |
| This Study |  |  | 213.79 [CI: 211.99, 215.59) |  |  |

| **CVT INCIDENCE ESTIMATES – ALL STUDIES** | | | |
| --- | --- | --- | --- |
| **Author**  **[date]** | **Design** | **Outcomes** | **Incidence CVT**  **Per 100,000** |
| Coutinho (2012)^1^ | Retrospective cross-sectional  Netherlands  Time: 2008-2010  Age: 18+ yrs | CVT:  ICD-9: 325, 437.6, 671.5, 437.8 | 1.32 [95% CI: 1.06–1.61].  2.78 women 31-50 |
| Devasagayam (2016)^2^ | Austrailia  Time: 2005-2011  Age: 19-84 | CVT: confirmed cases coding and/or neuroimaging  ICD-9: 325, 437.6, 671.5; *ICD-10*: I676. | 1.57 [CI: 1.29-1.90] |
| Otite (2020)^3^ | Hospital-based cohort study 2 states: FL & NY  Time: 2006-2014.  Age: 18+ | Cerebral Venous Thrombosis  ICD-9: 437.6, 325, 671.5x  ICD-10: I63.6, I67.6, O22.5X, O87.3, G08. | Crude:  2015: 1.84  2016: 2.03  Age+sex Stand:  2014: 2.02 [CI: 1.94-2.10]  2015: 1.87 [CI: 1.79-1.95]  2016: 2.00 [CI: 1.92-2.08] |
| Williame,  European Medicine Agency  (2021)^4^ | Claims data from across Europe | Cerebral Venous Thrombosis | Spain (BIFAP):  2017: 0.30 [CI: 0.21, 0.43]  2018: 0.31 [CI: 0.22, 0.45]  2019: 0.26 [CI: 0.17, 0.39]  Spain (SIDIAP)  2017: 0.24 [CI: 0.14, 0.41]  2018: 0.37 [CI: 0.24, 0.57]  2019: 0.39 [CI: 0.25, 0.60]  Italy (ARS):  2017: 1.20 [CI: 0.87, 1.66]  2018: 1.31 [CI: 0.96, 1.79]  2019: 1.51 [CI: 1.13, 2.01]  UK (CPRD):  2017: 0.14 [CI: 0.06, 0.34]  2018: 0.12 [CI: 0.05, 0.32]  2019: 0.15 [CI: 0.06, 0.36] |
| This Study |  | 6.37 [CI: 6.07-6.69] |  |
